# Effectiveness of 2024–2025 COVID-19 Vaccination Against COVID-19 Hospitalization and Severe In-Hospital Outcomes — IVY Network, 26 Hospitals, September 1, 2024–April 30, 2025

**DOI:** 10.1101/2025.08.29.25334612

**Authors:** Kevin C. Ma, Alexander Webber, Adam S. Lauring, Emily Bendall, Leigh K. Papalambros, Basmah Safdar, Adit A. Ginde, Ithan D. Peltan, Samuel M. Brown, Manjusha Gaglani, Shekhar Ghamande, Cristie Columbus, Nicholas M. Mohr, Kevin W. Gibbs, David N. Hager, Matthew E. Prekker, Michelle N. Gong, Amira Mohamed, Nicholas J. Johnson, Akram Khan, Catherine L. Hough, Abhijit Duggal, Jennifer G. Wilson, Nida Qadir, Steven Y. Chang, Christopher Mallow, Laurence W. Busse, Jennie H. Kwon, Matthew C. Exline, Ivana A. Vaughn, Mayur Ramesh, Jarrod M. Mosier, Aleda M. Leis, Estelle S. Harris, Adrienne Baughman, Sydney A. Cornelison, Paul W. Blair, Cassandra A. Johnson, Nathaniel M. Lewis, Sascha Ellington, Todd W. Rice, Carlos G. Grijalva, H. Keipp Talbot, Jonathan D. Casey, Natasha Halasa, James D. Chappell, Yuwei Zhu, Wesley H. Self, Fatimah S. Dawood, Diya Surie

## Abstract

**Importance:** As SARS-CoV-2 JN.1 lineage descendants continue to evolve, evaluating COVID-19 vaccine effectiveness (VE) against severe COVID-19 is necessary to inform vaccine composition updates.

**Objective:** To estimate effectiveness of 2024–2025 COVID-19 vaccines against COVID-19– associated hospitalizations and severe in-hospital outcomes overall and by time since dose (7– 89, 90–179, and ≥180 days), JN.1 descendant lineage (KP.3.1.1, XEC, LP.8.1), and spike mutations potentially associated with immune evasion.

**Design, setting, and participants:** This test-negative, case-control analysis included adult patients hospitalized during September 1, 2024–April 30, 2025 at 26 hospitals in 20 U.S. states. Cases presented with COVID-19–like illness and a positive SARS-CoV-2 nucleic acid or antigen test; controls had COVID-19–like illness but tested negative.

**Exposure:** Receipt of 2024–2025 COVID-19 vaccine ≥7 days before illness onset.

**Main Outcomes and Measures:** Main outcomes were COVID-19–associated hospitalization and severe in-hospital outcomes (supplemental oxygen therapy, acute respiratory failure, intensive care unit admission, invasive mechanical ventilation [IMV] or death). Logistic regression was used to estimate the odds of vaccination in cases and controls adjusting for demographics, clinical characteristics, and enrollment region. VE was estimated as (1 – adjusted odds ratio) x 100%.

**Results:** 1,888 COVID-19 cases (including 348 with KP.3.1.1, 218 with XEC, and 134 with LP.8.1 infections) and 6,605 controls were enrolled (median [IQR] age, 66 [54–76] years; 4,338 [51%] female). VE against COVID-19–associated hospitalization was 40% (95% CI, 27%–51%) and protection was sustained through 90–179 days after vaccination. VE was higher against the most severe outcome of IMV or death at 79% (95% CI, 55%–92%). VE was 49% (95% CI, 25%–67%) against hospitalization with KP.3.1.1, 34% (95% CI, 4%–56%) against XEC, and 24% (95% CI, -19% to 53%) against LP.8.1, with increasing median time since dose receipt due to sequential circulation patterns (60, 89, and 141 days, respectively). VE was similar against lineages with spike protein S31 deletion (41% [95% CI, 22%–56%]) and T22N and F59S substitutions (37% [95% CI, 9%–57%]).

**Conclusion and Relevance:** 2024–2025 COVID-19 vaccines provided additional protection against severe disease as multiple JN.1 descendant lineages circulated.

**Key points:** *Question:* What was estimated effectiveness of 2024–2025 COVID-19 vaccines against severe COVID-19 and did effectiveness vary by SARS-CoV-2 lineage or spike protein mutations?

*Findings:* Among 1,888 adult COVID-19 cases and 6,605 controls included in a test-negative, case-control analysis during September 2024–April 2025, vaccine effectiveness was 40% against hospitalization and 79% against invasive mechanical ventilation or death. Vaccine effectiveness was similar for KP.3.1.1 and XEC lineages and spike mutations potentially associated with immune evasion (S31 deletion, T22N and F59S substitutions).

*Meaning:* COVID-19 vaccines protected against severe disease during the 2024–2025 season when multiple JN.1 lineages evolved and circulated.

## Introduction

COVID-19 remains a public health threat with an estimated 320,000 to 480,000 hospitalizations and 37,000 to 56,000 deaths occurring nationally during the 2024–2025 season [1]. COVID-19 vaccination reduces the likelihood of severe COVID-19 [2], and timely estimates of vaccine effectiveness (VE) against new severe acute respiratory syndrome coronavirus 2 (SARS-CoV-2) variants can inform decisions on updates to COVID-19 vaccine composition [3]. In response to the shift in predominance from XBB to JN.1 lineages in January 2024, the U.S. Food and Drug Administration (FDA) approved updated 2024–2025 Moderna and Pfizer-BioNTech monovalent vaccines based on KP.2 and an updated Novavax vaccine based on JN.1 in August 2024 [4].

During the 2024–2025 COVID-19 season, circulating SARS-CoV-2 lineages remained JN.1 descendants with no major strain replacement event [5]. Instead, JN.1 descendant lineages repeatedly acquired spike protein substitutions and deletions in the N-terminal and receptor-binding domains that were associated with *in vitro* immune evasion [5–11]. JN.1 descendant lineages KP.3.1.1 and XEC contain 3 to 4 spike protein differences compared to KP.2 and were prevalent in U.S. genomic surveillance in late 2024; LP.8.1 contains 6 spike differences and increased starting early 2025 [5,12]. During September 2024 through early June 2025, receipt of a 2024–2025 COVID-19 vaccine was recommended for all persons in the U.S. aged ≥6 months [2,13]; vaccination coverage among adults participating in the National Immunization Survey reached 23% overall and 44% among adults aged ≥65 years [14].

We estimate effectiveness of 2024–2025 COVID-19 vaccines in preventing COVID-19-associated hospitalizations and severe in-hospital outcomes among US adults aged ≥18 years. Using whole-genome sequencing, we also characterize lineage- and mutation-specific VE against COVID-19-associated hospitalization.

## Methods

This analysis was conducted by the Investigating Respiratory Viruses in the Acutely Ill (IVY) Network, a multicenter, surveillance network comprising 26 hospitals in 20 US states. The IVY Network uses a test-negative, case-control design to assess VE; methods have been described previously [15–18]. Briefly, site personnel prospectively enrolled adult patients ≥18 years of age admitted to IVY Network hospitals who met a COVID-19–like illness case definition (Supplementary Methods) and received SARS-CoV-2 clinical testing. Enrolled patients were tested clinically for SARS-CoV-2 at the local hospital and additionally had nasal swab specimens collected and tested at a central laboratory at Vanderbilt University Medical Center for SARS-CoV-2, influenza viruses, respiratory syncytial virus (RSV), and human metapneumovirus (hMPV) using real-time reverse transcription–polymerase chain reaction (RT-PCR) (Supplementary Methods). Cases were defined by a positive test for SARS-CoV-2 in local or central laboratories for a specimen collected within 10 days of symptom onset and 3 days of hospital admission. Cases with known coinfections (i.e., influenza viruses, RSV, or hMPV) were excluded because COVID-19 vaccination would not be expected to prevent hospitalization caused by other respiratory viruses. Controls were defined by a negative test for SARS-CoV-2. Controls testing positive for influenza (all adults) or RSV (adults aged ≥60 years) were excluded due to correlation between COVID-19 and influenza or RSV vaccination behaviors, which can bias VE estimates [19–21]. Demographic and clinical data were collected through electronic medical record (EMR) review and patient or proxy interview. Data on race and ethnicity were collected because the association between COVID-19 case status and vaccination could vary by race or ethnicity.

This activity was reviewed by the CDC and each participating institution in the IVY Network, deemed public health surveillance and not research with waiver of participant informed consent, and was conducted consistent with applicable federal law and CDC policy [45 CFR part 46.102(l)(2), 21 CFR part 56; 42 USC §241(d); 5 USC §552a; 44 USC §3501 et seq]. This study is reported following the Strengthening the Reporting of Observational Studies in Epidemiology (STROBE) reporting guideline [22].

### Classification of Vaccination Status

Verification of COVID-19 vaccination status was performed using hospital EMRs, immunization information systems (IIS) (i.e., state or local vaccine registries), or patient or proxy interviews using a hierarchical approach. Evidence of vaccination from either EMRs or IIS were preferentially used even if interview data were available. If EMR/IIS data were unavailable, plausible interview data with known location and date of COVID-19 vaccine receipt were used [23]. Patients were classified into 2 vaccination groups: (1) those who received a 2024–2025 COVID-19 vaccine dose (either BNT162b2 [Pfizer-BioNTech], mRNA-1273 [Moderna], or NVX-CoV2705 [Novavax]) ≥7 days before illness onset and (2) those who did not receive a 2024– 2025 dose, comprising both patients who had received previous COVID-19 vaccine doses and patients who had never received a COVID-19 vaccine dose. We excluded patients if they received a 2024–2025 dose fewer than 7 days before illness onset or received more than one 2024–2025 dose.

### Sequencing Methods

SARS-CoV-2–positive specimens were sent to the University of Michigan (Ann Arbor, Michigan) for whole-genome sequencing to identify spike protein mutations and lineages. Sequencing was performed using the Oxford Nanopore Technologies Midnight protocol on a GridION instrument. Sequences were considered adequate for lineage identification if they had a Nextclade (version 3.4.1) genome coverage ≥80% and quality control status of “good” or “mediocre.” Lineages were defined from the following clades identified using Nextstrain nomenclature (representative Pango lineage shown in parentheses): 24A (JN.1), 24B (JN.1.11.1), 24C (KP.3), 24D (XDV.1), 24E (KP.3.1.1), 24F (XEC), 24G (KP.2.3), 24H (LF.7), 25A (LP.8.1), 25C (XFG). Spike amino acid substitutions and deletions were identified and parsed from Nextclade output.

### Severe In-Hospital Outcomes

The following severe in-hospital outcomes were characterized from hospital admission to the first of hospital discharge, patient death, or hospital day 28 (Supplementary Methods) [15]: (1) supplemental oxygen therapy (defined as supplemental oxygen at any flow rate and by any device for those not on chronic oxygen therapy, or with escalation of oxygen therapy for patients receiving chronic oxygen therapy), (2) acute respiratory failure treated with advanced respiratory support (defined as new receipt of high-flow nasal cannula, noninvasive ventilation, or invasive mechanical ventilation [IMV]), (3) intensive care unit (ICU) admission, and (4) a composite of IMV or death. To test the hypothesis that VE differed by outcome, *P-*values and 95% CIs for differences in VE against hospitalization versus VE against severe outcomes were calculated using bootstrapping with 10,000 replicates (Supplementary Methods).

### Statistical Methods

Descriptive comparisons were made using Pearson’s chi-squared test for categorical variables and the Mann-Whitney U test for continuous variables. Multivariable logistic regression was used to estimate the odds of 2024–2025 COVID-19 vaccination between cases and controls. Odds ratios were adjusted for age (continuous), sex (male, female), race/ethnicity (Hispanic or Latino, non-Hispanic Black, non-Hispanic White, non-Hispanic other race, unknown), US Department of Health and Human Services Region, admission date in biweekly intervals, and Charlson comorbidity index (0, 1–2, 3–4, 5–6, ≥7). VE against COVID-19–associated outcomes was calculated as (1 − adjusted odds ratio) × 100%. Estimates of VE were calculated separately for age group, immunocompromised status, time since vaccination strata (either 60- or 90-day windows), severe in-hospital outcomes, and cases with lineage (Nextstrain clade) or spike protein mutations. Statistical significance was indicated by a 2-sided *P*-value < 0.05. R (version 4.4.3; R Foundation for Statistical Computing) was used to conduct all analyses.

## Results

Among 8,493 adults included from 26 hospitals in the IVY Network during September 1, 2024, to April 30, 2025, 1,888 (22%) were COVID-19 cases and 6,605 (78%) were controls (Table 1, Supplementary Figures 1 and 2). Among included patients, median age was 66 years (interquartile range [IQR] = 54–76 years), 58% were non-Hispanic White, 51% were female, and median Charlson comorbidity index was 4 (IQR = 3–6) (Table 1). Controls had lower median age (65 years; IQR = 53–75 years) than cases (71 years; IQR = 59–80 years) (*P* < .001) and lower median Charlson comorbidity index (4; IQR = 2–6) than cases (5; IQR = 3–7) (*P* < .001). Distributions of sex, race and ethnicity, and enrollment site were similar (Table 1).

**Table 1.**
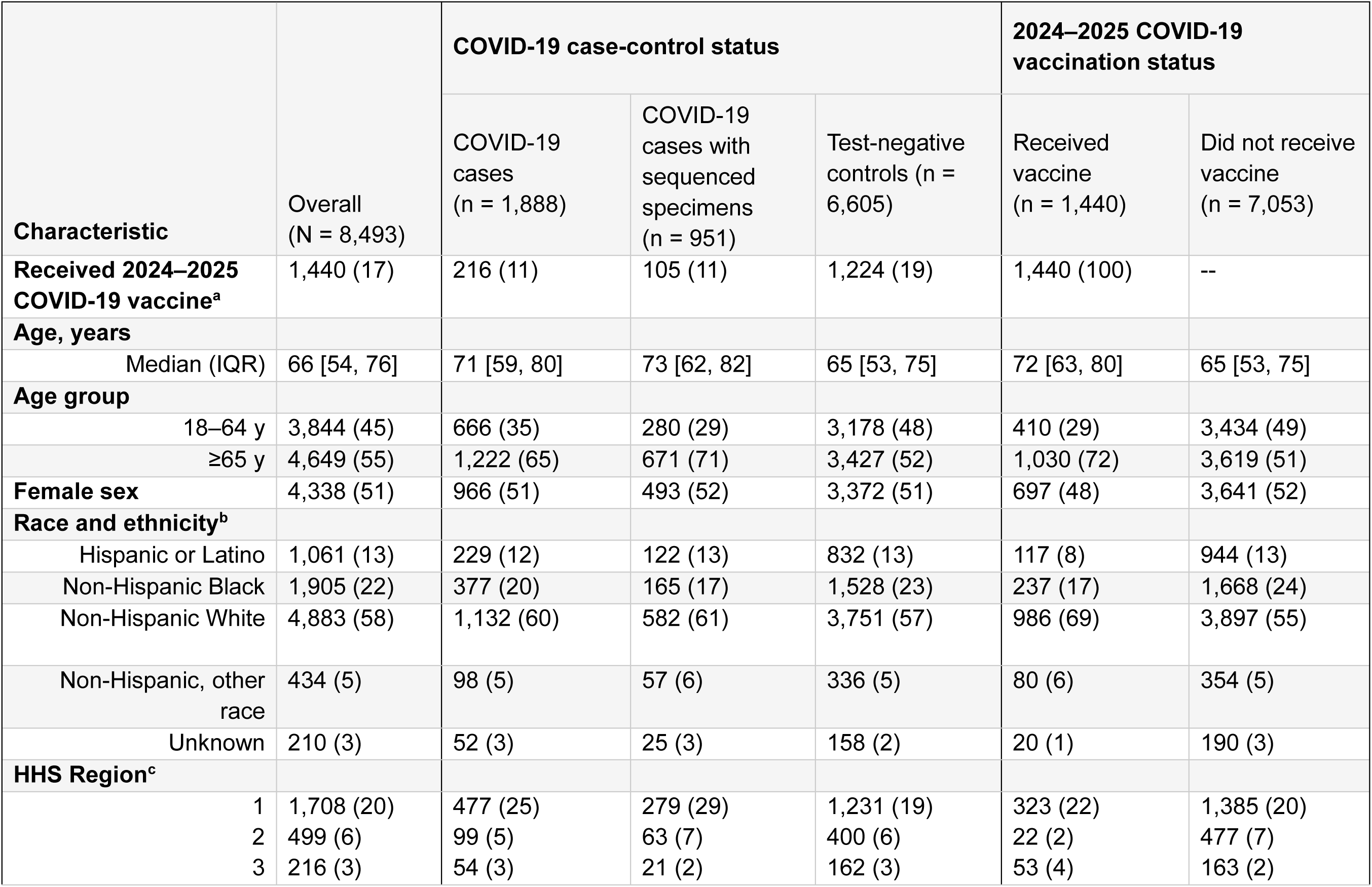
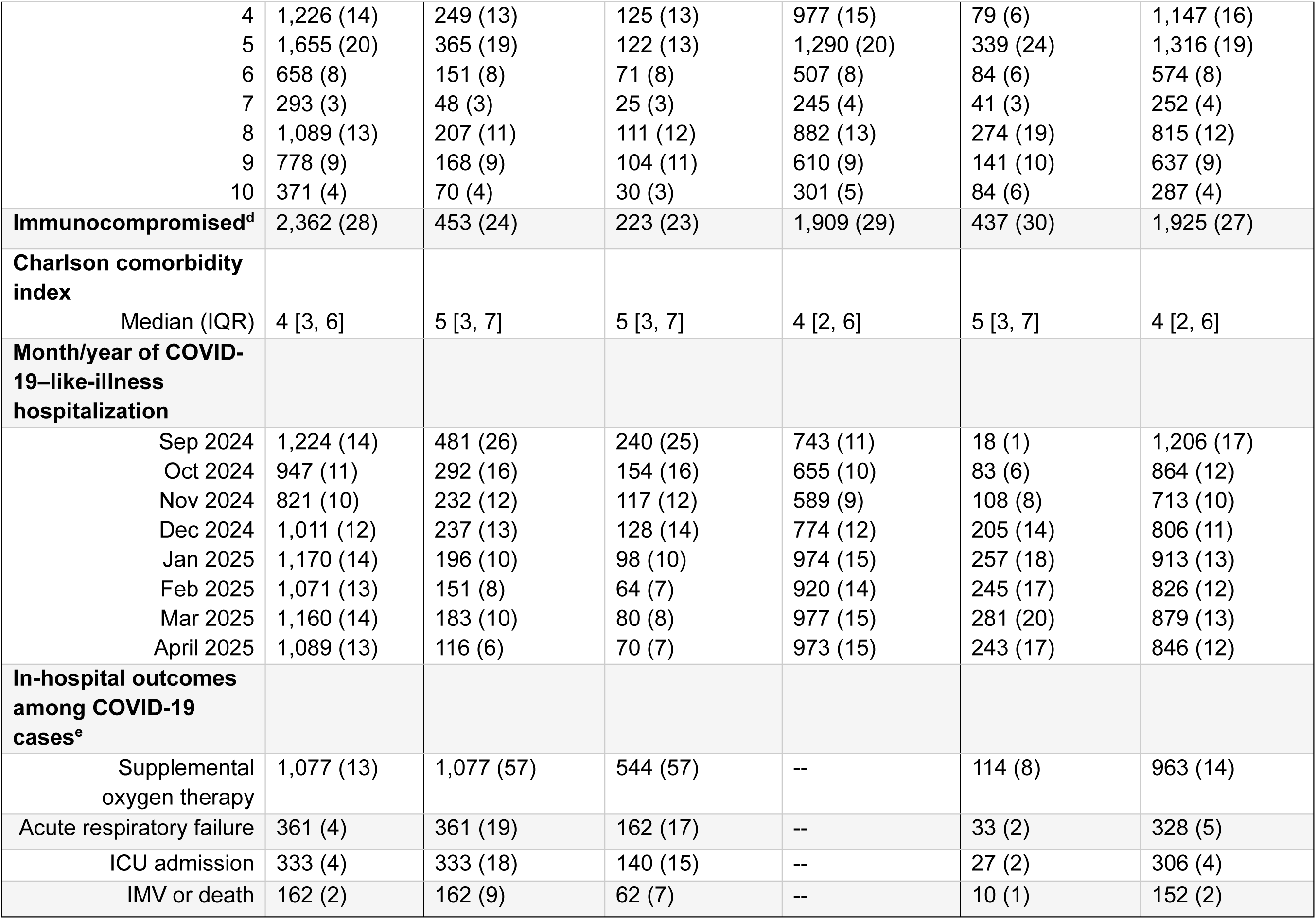
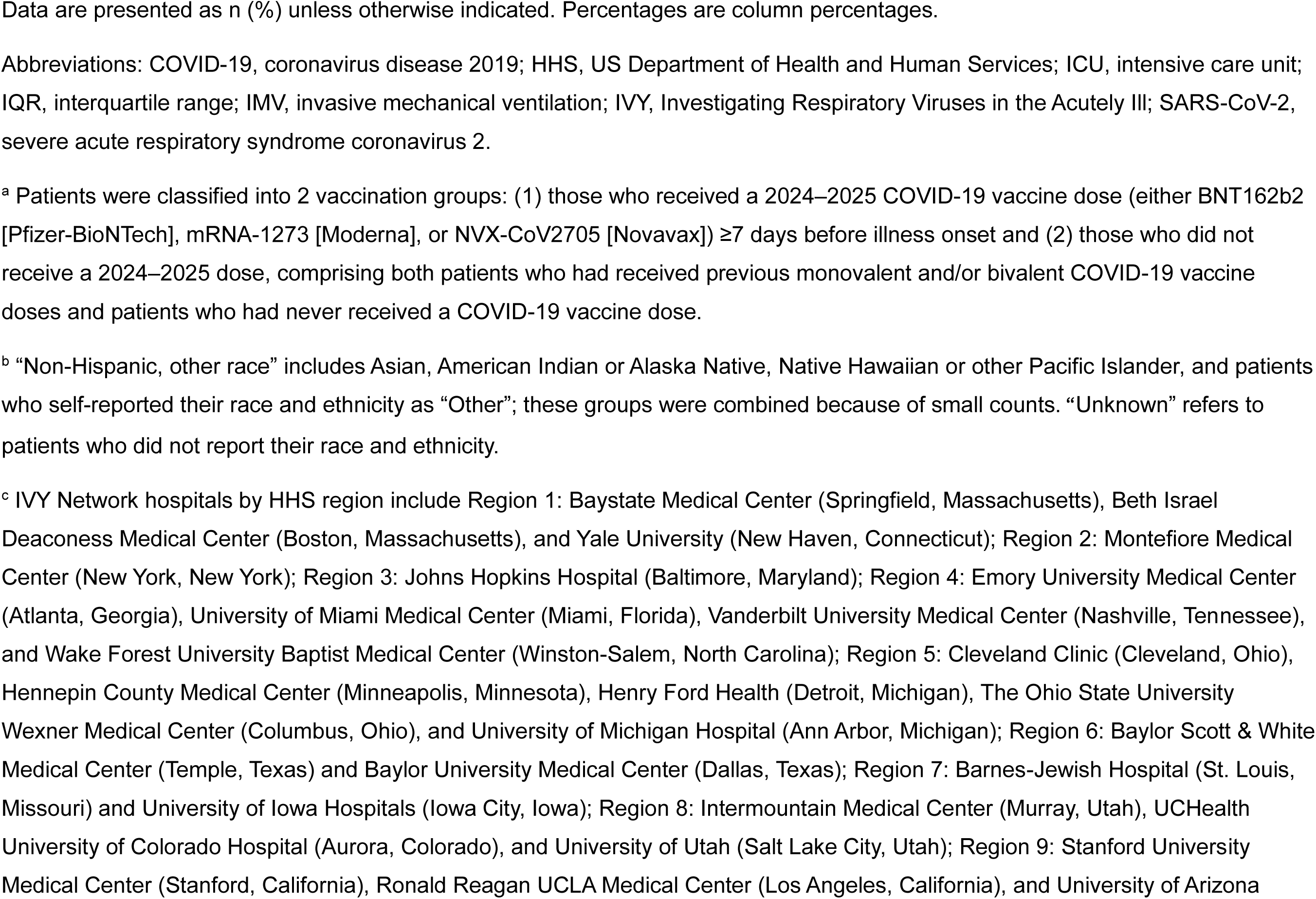
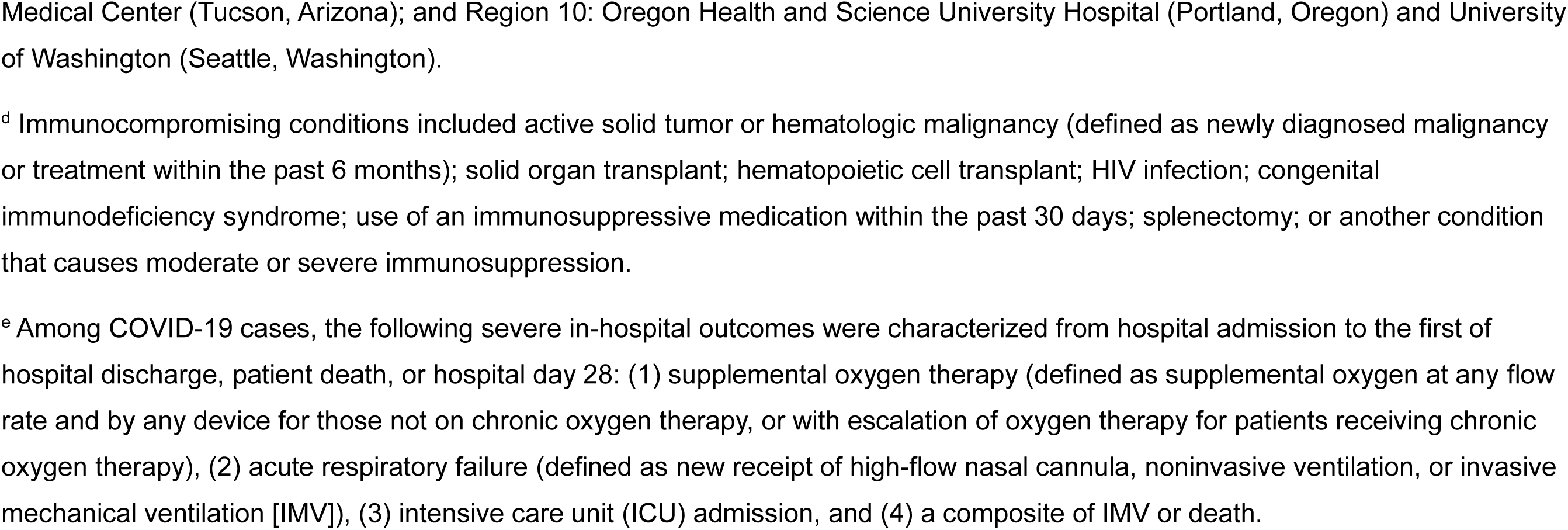
Characteristics of hospitalized adults aged ≥18 years with COVID-19-like illness by COVID-19 case-control and vaccination status – IVY Network, 26 hospitals, September 1, 2024–April 30, 2025.

A total of 216/1,888 (11%) cases and 1,224/6,605 (19%) controls received a 2024–2025 COVID-19 vaccine (Table 1). Weekly prevalence of 2024–2025 COVID-19 vaccination among controls increased from September 2024 to January 2025 before plateauing at approximately 20% (Supplementary Figure 3). Documentation of vaccination status was based on hospital EMR or vaccine registries where available (n = 1,274; 88%) and plausible patient or proxy interviews otherwise (n = 166; 12%). Among 1,251 patients with known product type information, 793 (63%) received BNT162b2 (Pfizer-BioNTech), 428 (34%) received mRNA-1273 (Moderna), and 30 (2%) received NVX-CoV2705 (Novavax).

### COVID-19 VE against hospitalization and severe in-hospital outcomes

Among 6,131 immunocompetent adults aged ≥18 years, overall effectiveness of 2024–2025 COVID-19 vaccine against COVID-19–associated hospitalization was 40% (95% CI, 27%–51%) with a median time since dose receipt of 80 days (IQR = 43–137 days) among cases and 108 days (IQR = 66–151 days) among controls. VE was 34% (95% CI, 14%–49%) 7–89 days and 52% (95% CI, 34%–65%) 90–179 days after vaccination (Figure 1), with similar trends by 60-day strata (Supplementary Figure 4). Among 3,450 immunocompetent adults aged ≥65 years, VE was 45% (95% CI, 31%–56%) overall, 44% (95% CI, 25%–59%) 7–89 days after vaccination, and 51% (95% CI, 31%–66%) 90–179 days after vaccination. Overall VE for immunocompromised adults aged ≥65 years (n = 1,199) was 36% (95% CI, 6%–57%) with a median time since dose receipt of 79 days (IQR = 49–131 days) among cases and 106 days (IQR = 56–153 days) among controls (Supplementary Figure 5).

**Figure 1.**
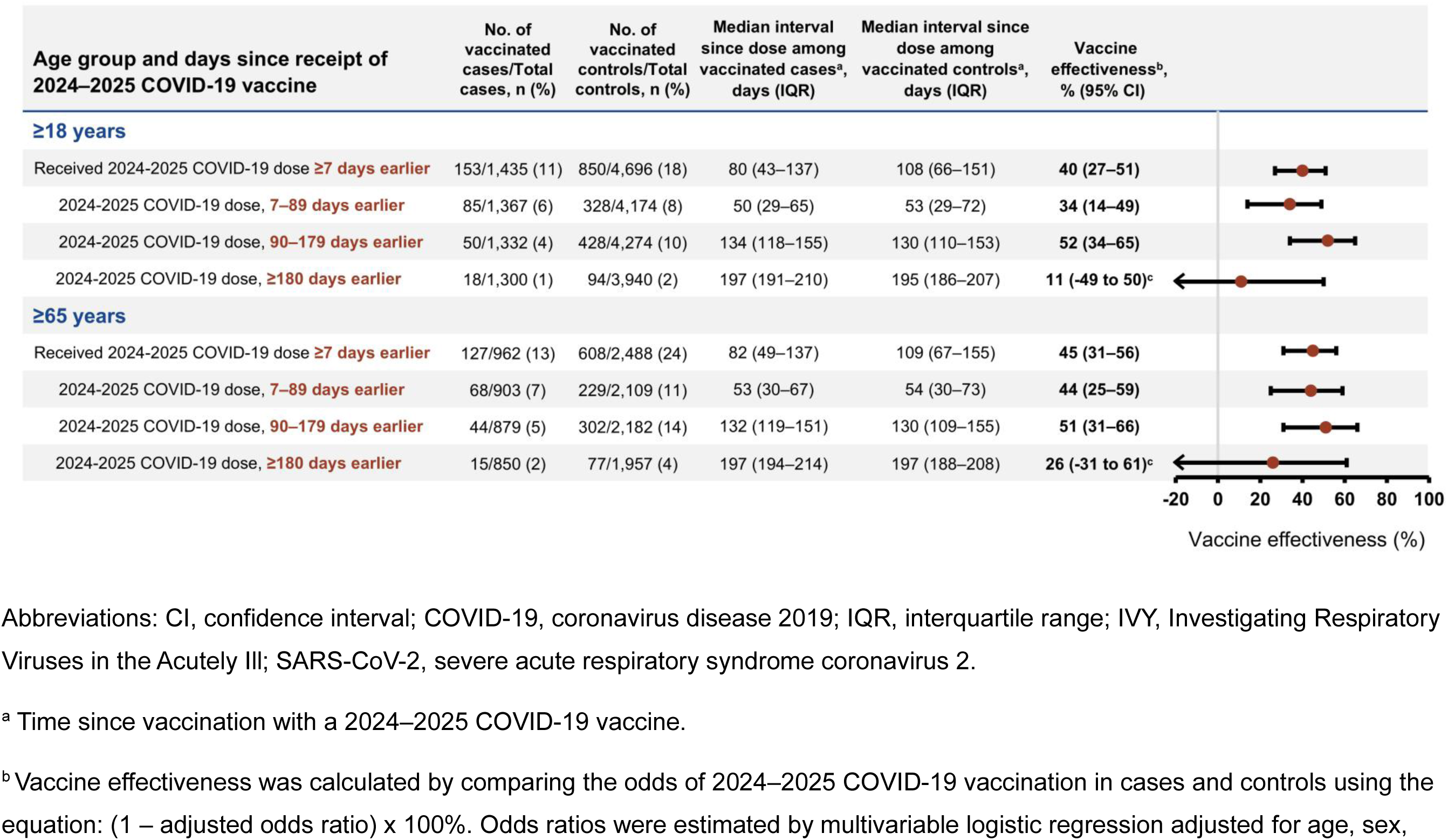
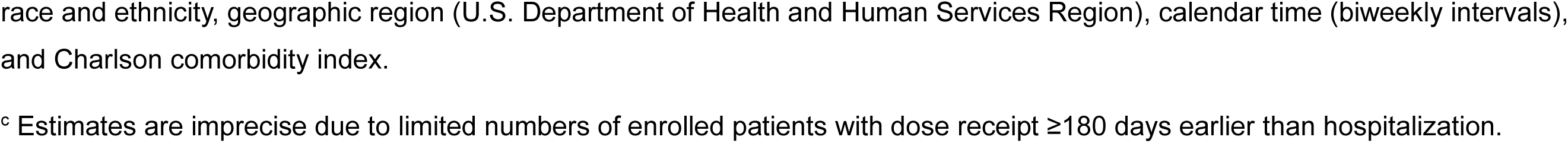
Effectiveness of 2024–2025 COVID-19 vaccine against COVID-19–associated hospitalization among immunocompetent adults by time since dose receipt and age group – IVY Network, 26 hospitals, September 1, 2024–April 30, 2025.

Among 1,888 COVID-19 cases, 1,077 (57%) received supplemental oxygen therapy, 361 (19%) experienced acute respiratory failure, 333 (18%) were admitted to the ICU, and 162 (9%) received IMV or died (Table 1). VE of a 2024–2025 dose for immunocompetent adults was 46% (95% CI, 31%–59%) against supplemental oxygen therapy, 49% (95% CI, 22%–68%) against acute respiratory failure, 60% (95% CI, 36%–77%) against ICU admission, and 79% (95% CI, 55%–92%) against receipt of IMV or death (Figure 2). Point estimates increased with more severe outcomes, and VE against IMV or death was significantly higher than VE against hospitalization (bootstrap *P* = 0.004; difference in VE estimates 39%, 95% bootstrap CI, 18%– 57%; Supplementary Table 1). Estimates were similar for immunocompetent adults aged ≥65 years, though VE was not significantly different between in-hospital outcomes and hospitalization (Figure 2, Supplementary Table 1).

**Figure 2.**
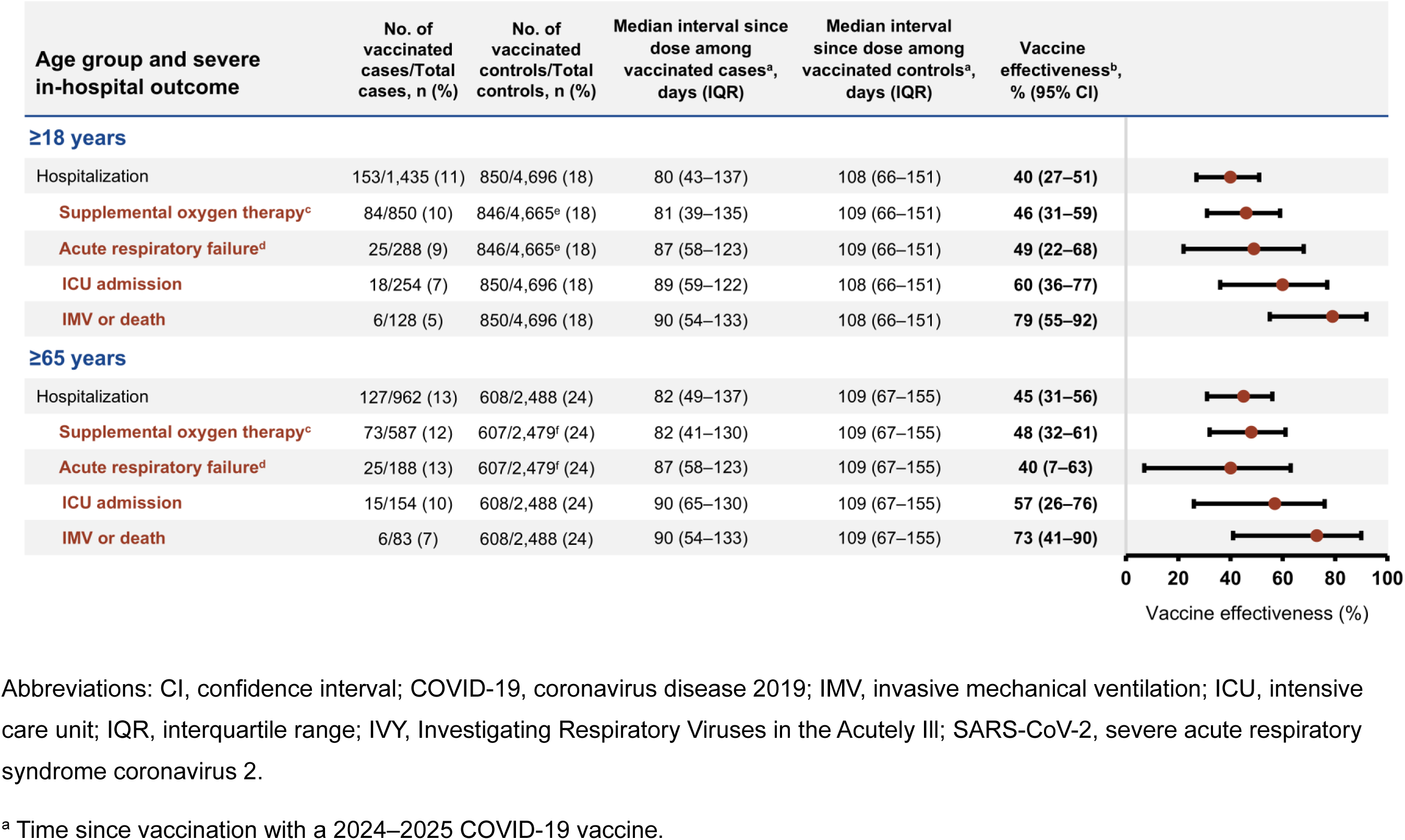
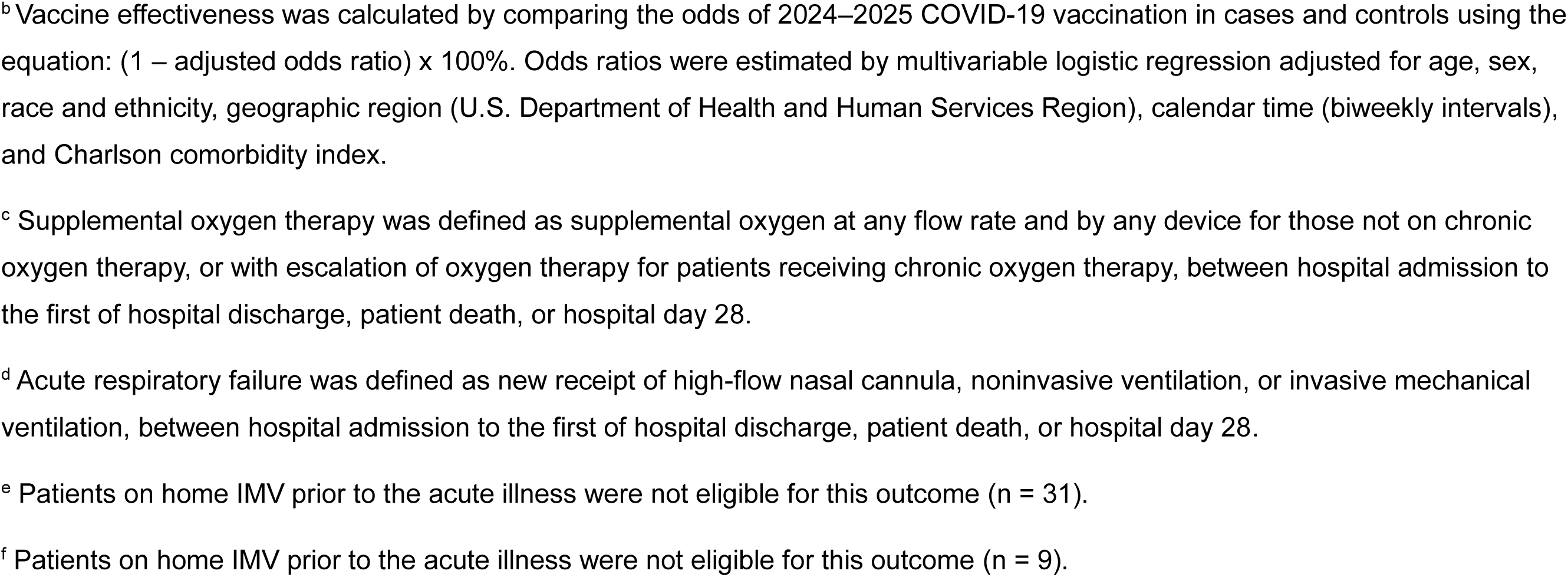
Effectiveness of 2024–2025 COVID-19 vaccine against COVID-19–associated severe in-hospital outcomes among immunocompetent adults by age group and outcome – IVY Network, 26 hospitals, September 1, 2024–April 30, 2025.

### COVID-19 VE by SARS-CoV-2 lineage and spike protein mutations

Identification of SARS-CoV-2 lineage through whole-genome sequencing was successful for 951 (50%) cases. Cases with sequencing data had similar distributions of clinical and demographic characteristics compared to all COVID-19 case-patients (Table 1). In a sensitivity analysis, VE by time since dose was similar for cases with sequencing data compared to all COVID-19 cases (Supplementary Figure 6). Among cases with sequencing, 348 (37%) had KP.3.1.1 lineage infection, 218 (23%) had XEC, 134 (14%) had LP.8.1, and the remainder (26%) were other JN.1 descendant lineages (Figure 3). KP.3.1.1 was most prevalent in September and October 2024, after which prevalence of XEC increased steadily from November 2024 to January 2025 before being displaced by LP.8.1 by February 2025. Nearly all circulating lineages contained either the S31 deletion (n = 602; 63%), T22N substitution (n = 290; 30%), or F59S substitution (n = 243; 26%) in the N-terminal domain of the spike protein (Supplementary Figure 7). The S31 deletion was more common among KP.3.1.1 (347/348), KP.2.3 (43/43), and LP.8.1 (134/134) and the T22N substitution was more common among XEC (217/218) and LF.7 (28/28) (Supplementary Table 2).

**Figure 3.**
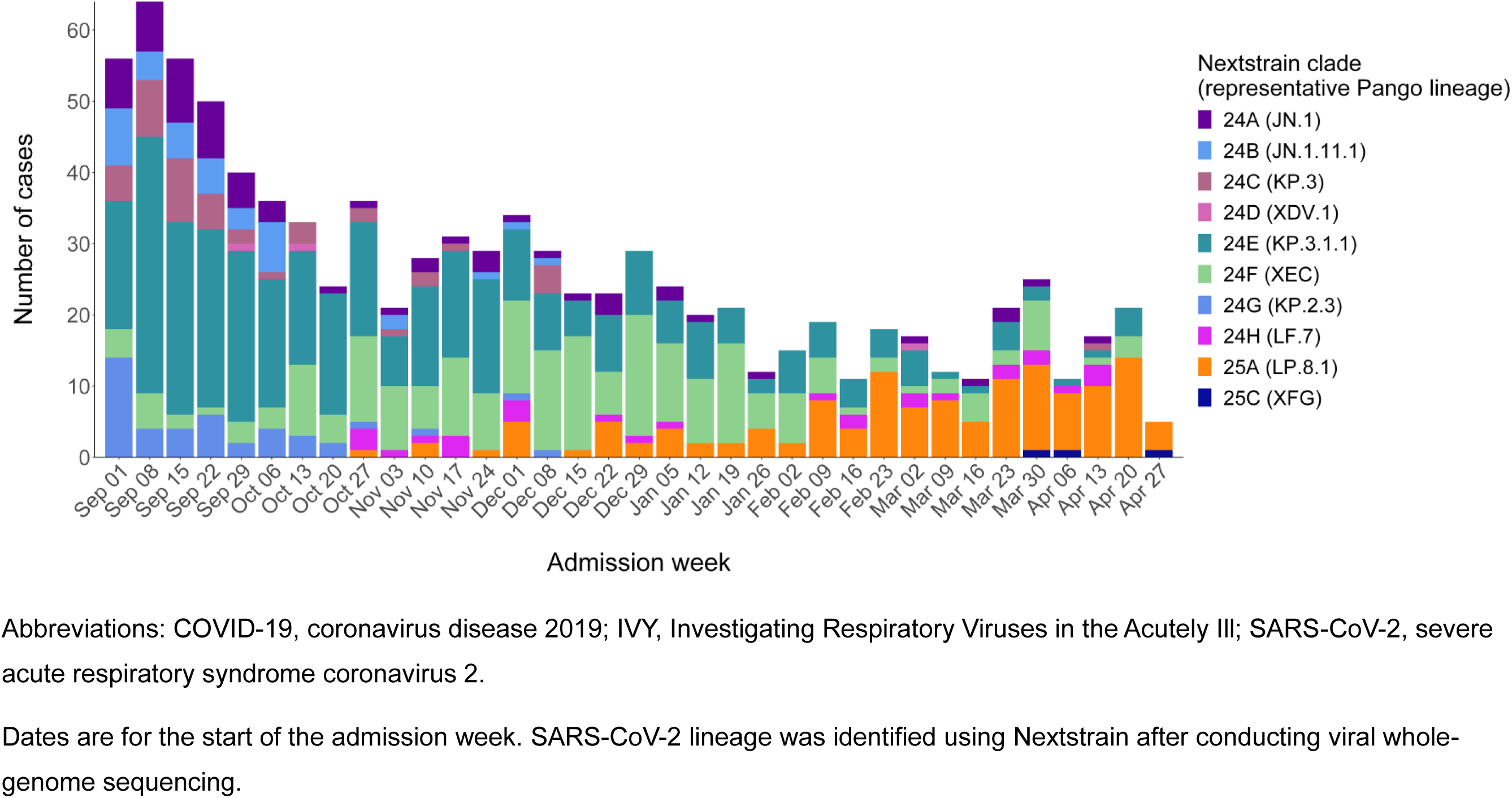
Number of COVID-19 cases by hospital admission week and SARS-CoV-2 lineage – IVY Network, 26 hospitals, September 1, 2024–April 30, 2025.

A total of 30 (9%) cases with KP.3.1.1, 35 (16%) with XEC, and 26 (19%) with LP.8.1 received a 2024–2025 COVID-19 vaccine dose. Among adults aged ≥18 years with available sequencing data (including both immunocompetent and immunocompromised patients), VE was 49% (95% CI, 25%–67%) against KP.3.1.1–associated hospitalization, 34% (95% CI, 4%–56%) against XEC–associated hospitalization, and 24% (95% CI, -19% to 53%) against LP.8.1–associated hospitalization (Figure 4). Due to sequential circulation (Figure 3), median time since dose receipt among cases was highest for LP.8.1 (141 days, IQR = 124–172 days) compared to XEC and KP.3.1.1 (*P* < 0.01), and higher for XEC (89 days, IQR = 55–122 days) compared to KP.3.1.1 (60 days, IQR = 35–79 days; *P =* 0.04). Restricting to the first 7–89 days following vaccination, VE was 43% (95% CI, 12%–65%) against KP.3.1.1–associated hospitalization and 48% (95% CI, 14%–70%) against XEC–associated hospitalization (Figure 4). VE was 41% (95% CI, 22%–56%) against SARS-CoV-2 strains with the S31 deletion and 37% (95% CI, 9%– 57%) against strains with the T22N and F59S substitutions, with similar time since dose receipt among cases (85 days, IQR = 50–143 and 89 days, IQR = 56–123, respectively) (Figure 4).

**Figure 4.**
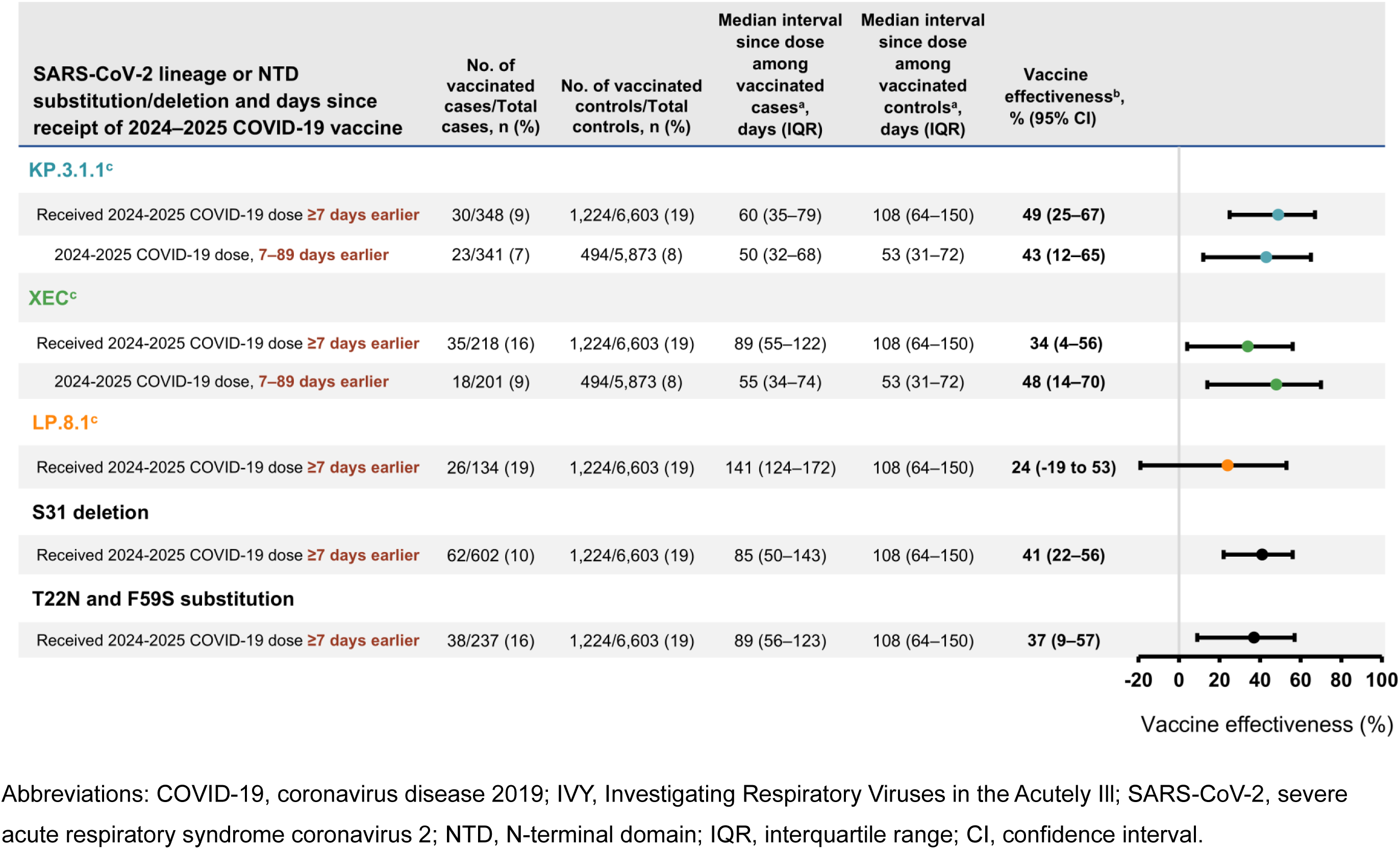
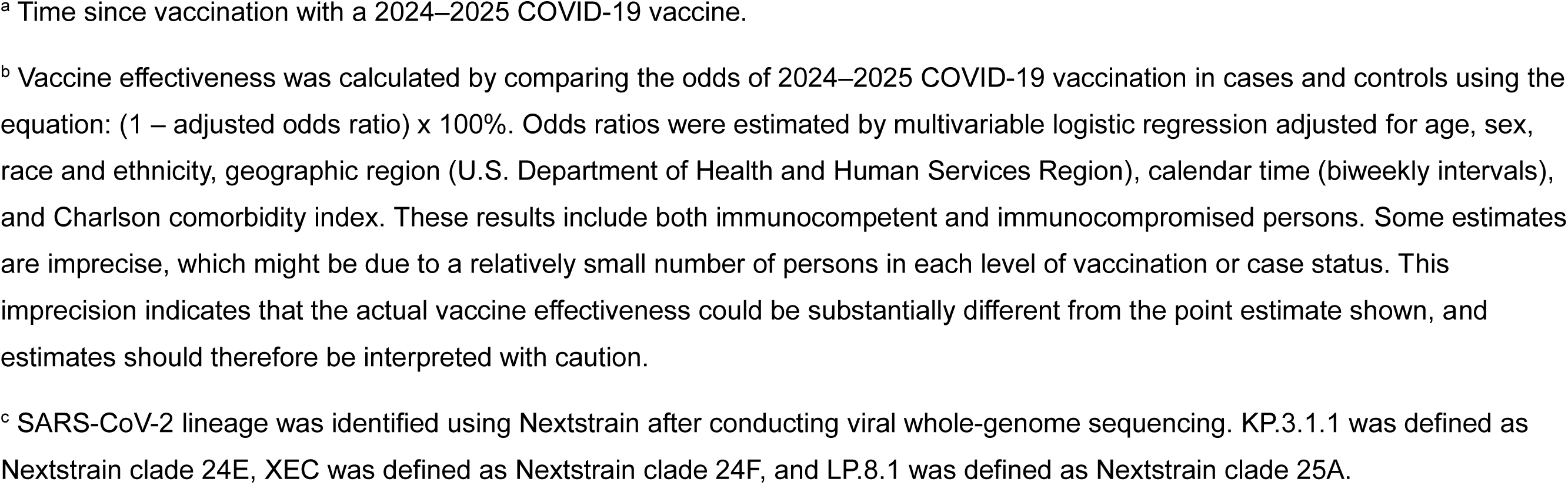
Effectiveness of 2024–2025 COVID-19 vaccine against hospitalization among adults aged ≥18 years by SARS-CoV-2 lineage and N-terminal domain substitutions or deletions – IVY Network, 26 hospitals, September 1, 2024–April 30, 2025.

## Discussion

In this multicenter US surveillance network, 2024–2025 COVID-19 vaccines were 40% effective against COVID-19-associated hospitalizations among immunocompetent adults enrolled September 2024 to April 2025. Protection was sustained until at least 3–6 months following vaccine receipt. Effectiveness was highest against the most severe outcomes of IMV or death (79%), consistent with earlier reports during the COVID-19 pandemic [16,24–29]. VE among all adults was similar against KP.3.1.1- and XEC-associated hospitalization in the first 3 months after vaccination (43% and 48%, respectively). VE was lower against LP.8.1–associated hospitalization (24%) with 95% CIs that overlapped the null, but estimates were imprecise and time since vaccination was higher compared to other lineages. VE was similar against variants with spike S31 deletions or T22N/F59S substitutions (41% and 37%, respectively). Taken together, these findings demonstrate JN.1/KP.2-based COVID-19 vaccines were effective against multiple JN.1 descendent lineages that emerged sequentially and co-circulated during the 2024–2025 season.

Our findings demonstrating added protection from 2024–2025 COVID-19 vaccines against hospitalization are concordant with other US estimates, including two electronic health record-based networks reporting VE of 68% and 45% a median of 30 and 53 days following vaccination, respectively [30,31]. Our estimates are also consistent with VE of JN.1-based vaccines from the United Kingdom (43%) [32], Denmark (85% against hospitalization, 96% against death) [33], and a multinational outpatient European network (66% against medically attended illness) [34]. We observed a lower VE point estimate 7–89 days compared with 90– 179 days after vaccination, similar to findings from the United Kingdom [32]. This may have been caused by transient increases in population immunity from increased SARS-CoV-2 circulation late summer 2024 just prior to availability of 2024–2025 COVID-19 vaccines.

Increases in population immunity can result in lower estimates of COVID-19 VE because VE is a measure of incremental protection beyond prior vaccination or infection [35,36]. Finally, few studies have evaluated protection against severe in-hospital outcomes this season, and our finding of higher VE against COVID-19–associated IMV or death demonstrates the importance of recent vaccination to prevent the most severe outcomes following infection.

Few observational studies have evaluated lineage-specific COVID-19 VE during the 2024–2025 season. One challenge is that several JN.1 descendants co-circulated without clearly defined periods of predominance (Figure 3). In such a scenario, lineage-specific VE estimates based on calendar time to delineate predominance periods could be biased due to lineage misclassification error [37], and whole-genome sequencing of patient specimens may be the only reliable approach for estimation. A study using whole-genome sequencing and national registry data in Denmark found similar protection against KP.3.1.1, XEC, and overall COVID-19– associated hospitalizations from JN.1-based vaccines [33]. Immunogenicity studies have also found strong increases in neutralizing antibody responses to KP.3.1.1 and XEC following JN.1/ KP.2-based booster administration [38–40]. We were unable to distinguish the effect of natural waning from variant-mediated immune escape for LP.8.1 due to limited sample sizes, but antigenic cartography data suggest that LP.8.1 is similar to other JN.1 descendants [41,42], and JN.1/KP.2-based vaccination yields similar antibody titers against LP.8.1, KP.3.1.1, and XEC in some cohorts [42–45].

N-terminal domain mutations including the S31 deletion (present in KP.3.1.1, LP.8.1, and other lineages) and T22N and F59S substitutions (present in XEC) are a novel occurrence for SARS-CoV-2 and may confer fitness advantages through introduction of glycosylation sites, which can disrupt binding of N-terminal domain-binding antibodies [6,7,9–11]. The S31 deletion and F59S substitution have been hypothesized to also induce conformational changes in SARS-CoV-2 spike protein that could affect antibody binding to the receptor binding-domain [10]. Here, we demonstrate these N-terminal mutations were prevalent among patients hospitalized with COVID-19 and that 2024–2025 COVID-19 vaccines provided similar protection against lineages with S31 deletion versus T22N and F59S substitutions [9].

This analysis is subject to several limitations. First, the sequential timing of KP.3.1.1, XEC, and LP.8.1 circulation correlated with increasing time since vaccination, which made it challenging to differentiate effects of natural immunologic waning versus variant immune evasion, particularly for LP.8.1. Therefore, it is unclear if the lower point estimate for LP.8.1 was due to genetic changes or increased time since dose. Second, decreasing counts of COVID-19 hospitalizations in spring 2025 precluded precise estimation of VE beyond 180 days after vaccine receipt. Third, this analysis did not adjust for prior SARS-CoV-2 infection, which can influence estimates of VE [35]. Fourth, while we attempted to sequence specimens from all cases, we were unable to identify lineage in approximately half of specimens potentially due to low viral load or viral degradation. Exclusion of these patients did not appear to substantially bias VE (Supplementary Figure 5) [46]. Finally, although analyses were adjusted for some relevant confounders, residual confounding may remain.

Our results demonstrate 2024–2025 COVID-19 vaccines provided protection against COVID-19 hospitalization and severe in-hospital outcomes and were effective against multiple JN.1 descendants. During a season without major antigenic changes to circulating SARS-CoV-2 viruses, we found sustained protection from COVID-19 vaccines through at least 90–179 days after vaccination. In June 2025, the US Food and Drug Administration recommended monovalent JN.1 lineage-based COVID-19 vaccines, preferentially using the LP.8.1 strain, for 2025–2026 COVID-19 vaccine formulations [47]. Monitoring COVID-19 VE, including stratifying by SARS-CoV-2 lineage and spike protein mutations, remains important to guide COVID-19 vaccine composition and recommendations [3].

## Data Availability

No additional data available.

## Acknowledgements

Ryan Wiegand, PhD (CDC), and Gordana Derado, PhD (CDC) for statistical review. Benjamin Silk, PhD (CDC) for scientific review.

## Disclaimer

The findings, conclusions, and views expressed in this presentation are those of the authors and do not necessarily represent the official position of the Centers for Disease Control and Prevention (CDC).

