## Supplementary Materials for "Effectiveness of 2024–2025 COVID-19 Vaccination Against COVID-19 Hospitalization and Severe In-Hospital Outcomes — IVY Network, 26 Hospitals, September 1, 2024–April 30, 2025"

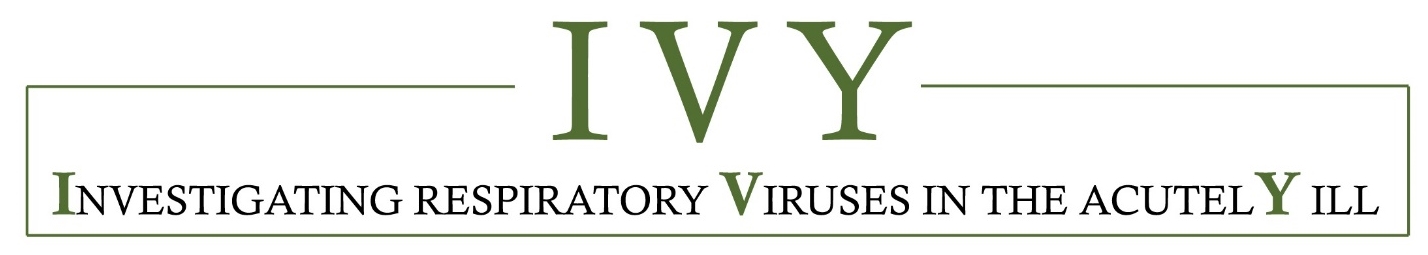

**Supplementary Materials**

Effectiveness of 2024–2025 COVID-19 Vaccination Against COVID-19 Hospitalization and Severe In-Hospital Outcomes — IVY Network, 26 Hospitals, September 1, 2024–April 30, 2025

Investigators and Collaborators………………………………………………………..…………………3

Supplementary Methods ………………………………………………………………….………………6

1. COVID-19 case definition……………..……………………………………………………..6
2. Severe in-hospital outcome definitions……………………………………………………..6
3. Laboratory testing methods..……………………………………………...…….…………..8
4. Analytic methods…………………………………………………………………………….10
5. Supplementary references...……………………………………………………………….12

Supplementary Tables and Figures..……………………………………………………………..…….13

Supplementary Table 1. Bootstrap-based *P*-values and 95% confidence intervals for differences in the effectiveness of 2024–2025 COVID-19 vaccine against COVID-19–associated hospitalization versus in-hospital severe outcomes...………………………….13

Supplementary Table 2. Counts of spike protein N-terminal domain substitutions or deletions by Nextstrain clades ………………………...………………………………………14

Supplementary Figure 1. Participant flow diagram and analytic cohorts………...………..15

Supplementary Figure 2. Numbers of COVID-19 cases and test-negative controls by admission week …………….………………………………………………………....………..16

Supplementary Figure 3. 2024–2025 COVID-19 vaccination coverage by admission week among test-negative controls………………………………………………………….……….17

Supplementary Figure 4. Effectiveness of 2024–2025 COVID-19 vaccine against COVID-19–associated hospitalization among immunocompetent adults by time since dose receipt (60-day windows) and age group……………………………………………………..18

Supplementary Figure 5. Effectiveness of 2024–2025 COVID-19 vaccine against COVID-19–associated hospitalization among immunocompromised adults by age group..……..20

Supplementary Figure 6. Effectiveness of 2024–2025 COVID-19 vaccine against COVID-19–associated hospitalization among cases with SARS-CoV-2 lineage successfully identified through whole-genome sequencing and among all cases by time since dose receipt …………………………………………..……………………..……………..…………..21

Supplementary Figure 7. Number of COVID-19 cases by hospital admission week and SARS-CoV-2 spike protein N-terminal domain substitutions and deletions………...…….23

**IVY Network Group**

Investigators and collaborators of the Investigating Respiratory Viruses in the Acutely Ill (IVY) Network are listed below.

**Baylor, Scott and White, Temple, Texas**

Manjusha Gaglani, Shekhar Ghamande, Tresa McNeal, Nicole Calhoun, Jocelyn Cravens, Kempapura Murthy, Leah Odame-Bamfo, Spencer Rose, Michael Smith, Barbara Hairston, Amanda McKillop, Victoria Harkins Walston

**Baylor, Scott and White, Dallas, Texas**

Manjusha Gaglani, Cristie Columbus, Robert L. Gottlieb, Catherine Raver, Sydney Buehrig, Priyanka Rana, Ashley Bychkowski, Symone Dunkley, Denisse Mariscal, Tammy Fisher, Daniela Gonzalez, Therissa Grefsrud, Mariana Hurutado-Rodriguez, Gabriela Perez

**Baystate Medical Center, Springfield, Massachusetts**

Jay Steingrub, Lesley De Souza, Scott Ouellette, Cynthia Kardos, Rae Lynn Defeo

**Beth Israel Medical Center, Boston Massachusetts**

Nathan I. Shapiro, Michael Bolstad, Brianna Coviello, Robert Ciottone, Arnaldo Devilla, Ana Grafals, Conor Higgins, Carlo Ottanelli, Kimberly Redman, Douglas Scaffidi, Alexander Weingart

**Centers for Disease Control and Prevention, Atlanta, Georgia**

Diya Surie, Kevin C. Ma, Alexander Webber, Fatimah S. Dawood, Nathaniel M. Lewis, Sascha Ellington

**Cleveland Clinic, Cleveland, Ohio**

Omar Mehkri, Megan Mitchell, Zachary Griffith, Connery Brennan, Kiran Ashok, Bryan Poynter, Abhijit Duggal

**Emory University, Atlanta, Georgia**

Laurence Busse, William Bender, Caitlin ten Lohuis

**Hennepin County Medical Center, Minneapolis, Minnesota**

Matthew Prekker, Laurynn Giles, Mary O'Rourke, Anne Frosch, Audrey Hendrickson, Julianna Prohofsky, Kowsar Hurreh, Joanna Kuo

**Henry Ford Health, Detroit, Michigan**

Ivana A Vaughn, Gina Maki, Mayur Ramesh, Jaskiran Bansal, Michael Garcia, Alycia Lilla, Catherine McKeon, Maria Santana-Garces, Tanisha Shack, Sindhuja Koneru, Shruti Tirumala

**Intermountain Medical Center, Murray, Utah**

Ithan D. Peltan, Samuel M. Brown, Joslyn Bassett, Shandi Poulson, Vineela Thumma

**Johns Hopkins University, Baltimore, Maryland**

David N. Hager, Minh Phan, Safa Saeed

**Montefiore Medical Center, Bronx, New York**

Michelle Gong, Amira Mohamed

**Ohio State Medical Center, Columbus, Ohio**

Sarah Karow, Maryiam Khan, Gabrielle Swoope, Brooke Lee, Amane Rasul, Kirstina Luikart, Tyler Haley, Amer Charif, Thomas Sturges, Ibrahim Abu Hammad, Rashil Madan

**Oregon Health and Sciences University, Portland, Oregon**

Jenny Chan, Connie Tran, Davika Dige, Anabelle Blue, Amy Segura, Riya Matthew, Adrian Hernandez, Emily Tribbett, Genesis Briceno

**Stanford University, Stanford, California**

Grace Kyin-Ye Tam, Vanessa Pitre, Samantha Ferguson, Cynthia Perez, Alexandra June Gordon, Leonard Basobas, Lily Lau

**University of Arizona, Tucson, Arizona**

Cameron Hypes, Karen Lutrick, Beth Salvagio Campbell

**University of California-Los Angeles, Los Angeles, California**

Cody Tran, Sukantha Chandrasekaran, Omai Garner

**University of Colorado, Aurora, Colorado**

Adit A. Ginde, Samantha Simon, Amanda Martinez, Amy Sullivan, Laura Aguilar-Marquez, Erika Alor, Yvette Evans, Jacob Rademacher

**University of Iowa, Iowa City, Iowa**

Nicholas Mohr, Anne Zepeski, Paul Nassar, Noble Briggs, Jacob Hampton, Cathy Fairfield

**University of Miami, Miami, Florida**

Chris Mallow, Carolina Rivas

**University of Michigan, Ann Arbor, Michigan**

Weronika Damek Valvano, Anne Kaniclides, Aleda Leis, Rebecca Fong, Mildred Wallace, Chiraag Balsara, Rachel Truson, Regina Lehmann, Abigail Carolan, Izza Imran, Jozyan Ujmaya, William J. Fitzsimmons

**University of Utah, Salt Lake City, Utah**

Rylie McBride, Bryce Bosworth, Bryce Heslop

**University of Washington, Seattle, Washington**

Joshua Acidera, Maile Mckeown, Leenay Coughlin, Frances Nagore, Dylan Clark

**Vanderbilt University Medical Center, Nashville, Tennessee**

Wesley Self, Yuwei Zhu, Cassandra Johnson, Adrienne Baughman, James D. Chappell, Natasha Halasa, Carlos G. Grijalva, Paul W. Blair, Jonathan D. Casey, Karen F. Miller, Jakea Johnson, Kelsey N. Womack, Jillian Rhoads, Colleen Ratcliff, Sydney Cornelison, Ine Sohn, Cara Lwin, and staff of the Vanderbilt Infection Surveillance and Prevention Laboratory (Julio Angulo, Stacy Batey, Shanice Cummings, Abby Fink, Claudia Guevara, Jennifer Luther, Rendie McHenry, Bryan Peterson, Neekar Rashid, Wanderson Rezende, Kalee Rumfelt, and Laura Short).

**Wake Forest University, Winston-Salem, North Carolina**

Kevin Gibbs, Hannah Strait

**Washington University, St. Louis, Missouri**

Jennie Kwon, Bijal Parikh, Carleigh Samuels, Lucy Vogt, Caroline O'Neil, Alyssa Valencia, Francesca Yerbic, Olivia Arter, Akshay Saluja, Elianora Ovchiyan, Sachina Mensah, Kim Vu, David McDonald, Regine Burton

**Yale University, New Haven, Connecticut**

Anirudh Goyal, Ivan Valesquez, Arda Yigitkanli, Kimberly Manchester

**Supplementary Methods**

**1. COVID-19 case definition**

COVID-19–like illness was defined as one or more of the following: fever, cough, shortness of breath, new or worsening findings on chest imaging consistent with pneumonia, or hypoxemia (defined as oxygen saturation <92% or supplemental oxygen use for patients without chronic oxygen needs, or escalation of oxygen therapy for patients on chronic supplemental oxygen).

**2. Severe in-hospital outcomes**

Clinical severity of COVID-19 cases was characterized using the following severe in-hospital outcomes occurring from hospital presentation to hospital discharge, patient death, or hospital day 28:

i) COVID-19-associated supplemental oxygen therapy

ii) COVID-19-associated acute respiratory failure

iii) COVID-19-associated intensive care unit (ICU) admission

iv) COVID-19-associated invasive mechanical ventilation (IMV) or death

i) COVID-19-associated supplemental oxygen therapy

Patients who met the definition of COVID-19-associated supplemental oxygen therapy either required supplemental oxygen therapy at any time during the hospitalization through day 28 for those not on chronic oxygen or, for patients on chronic supplemental oxygen (**Table**), required an escalation in respiratory support. Supplemental oxygen therapy could be delivered at any flow rate and by any device; this included standard flow oxygen (flow rate <30 liters/minute), high-flow nasal cannula (HFNC), non-invasive ventilation (NIV), and IMV. Patients on home IMV prior to the acute illness were not eligible for this outcome.

| Classification of in-hospital respiratory outcome based on type of oxygen or respiratory support used chronically (before illness onset) and highest level received through hospital day 28 | | | | |
| --- | --- | --- | --- | --- |
| **Chronic pre-illness oxygen use** | **Oxygen use during hospital course (highest support)** | **Is the patient eligible for this outcome?** | | |
|  |  | **Supplemental**  **oxygen therapy** | **Acute respiratory failure** | **Invasive mechanical ventilation** |
| No oxygen use | Standard flow oxygen | Yes | No | No |
|  | High-flow nasal cannula (HFNC) | Yes | Yes | No |
|  | NIV | Yes | Yes | No |
|  | IMV | Yes | Yes | Yes |
| Standard flow oxygen | Standard flow oxygen | No | No | No |
|  | HFNC | Yes | Yes | No |
|  | NIV | Yes | Yes | No |
|  | IMV | Yes | Yes | Yes |
| Non-invasive mechanical ventilation (NIV) | Standard flow oxygen | No | No | No |
|  | HFNC | No | No | No |
|  | NIV | No | No | No |
|  | IMV | Yes | Yes | Yes |
| Invasive mechanical ventilation (IMV) | Standard flow oxygen | No | No | No |
|  | HFNC | No | No | No |
|  | NIV | No | No | No |
|  | IMV | No | No | No |

ii) COVID-19-associated acute respiratory failure

Patients were classified as having COVID-19-associated acute respiratory failure treated with advanced respiratory support if they received any of the following during the hospitalization through day 28: HFNC, NIV, or IMV. HFNC was defined as a supplemental oxygen flow rate ≥30 liters per minute. NIV included both continuous positive airway pressure (CPAP) and bilevel positive airway pressure (BiPAP) delivered through a mask. Patients were classified as having NIV use if NIV was received for therapy of the acute illness and not only for treatment of sleep apnea. IMV was defined as positive pressure administered through an endotracheal tube or tracheostomy tube. Patients on home NIV before the acute illness met criteria for this outcome if they had escalation of respiratory support to IMV in the hospital. Patients on home IMV prior to the acute illness were not eligible for this outcome.

iii) COVID-19-associated intensive care unit (ICU) admission

Patients were classified as having COVID-19-associated ICU admission if they received care in an ICU for any duration of time during the hospitalization through day 28.

iv) COVID-19-associated IMV or death

Patients were classified as having COVID-19-associated IMV or death if they received IMV or died during the hospitalization through day 28. IMV was defined as positive pressure administered through an endotracheal tube or tracheostomy tube. Patients on home IMV prior to the acute illness could not meet the COVID-19-associated IMV or death outcome through receipt of in-hospital IMV.

**2. Laboratory Testing Methods**

At the time of participant enrollment, a nasal swab specimen was obtained via a fresh swabbing procedure or collection of a residual aliquot in the clinical laboratory. Specimens were stored at -80^o^C at the enrolling site and shipped to Vanderbilt University Medical Center. Real-time reverse transcription polymerase chain reaction (RT-PCR) testing for SARS-CoV-2, influenza, and RSV was completed at Vanderbilt. Specimens that tested positive for SARS-CoV-2 were submitted to the University of Michigan for viral whole-genome sequencing.

*SARS-CoV-2 detection by RT-PCR*

Total nucleic acid extract from 100 microliters of upper respiratory specimen collected in viral transport medium was prepared using the MagNA Pure LC Total Nucleic Acid Isolation Kit (Roche Molecular Systems, Pleasanton, CA) and MagNA Pure 96 automated extraction platform (Roche) or QIAamp 96 Virus QiaCube HT kit (Qiagen, Germantown, MD) and QiaCube HT automated extraction system (Qiagen). Extracts (100 microliters eluate volume) were tested by RT-PCR using the QuantStudio 3, QuantStudio 5, or QuantStudio 6 Real-Time PCR System (Applied Biosystems, Waltham, MA) for SARS-CoV-2 nucleocapsid (N)-gene N1 and N2 targets and for the human RNase P (RNP) gene with TaqPath 1-Step RT-qPCR Master Mix CG (Applied Biosystems) according to the *CDC 2019-Novel Coronavirus (2019-nCoV) Real-Time RT-PCR Diagnostic Panel* protocol (<https://www.fda.gov/media/134922/download>). Qualitative result assignment of *positive*, *not detected*, *inconclusive*, or *invalid specimen* for SARS-CoV-2 RNA was based on the pattern of N1, N2, and RNP Ct values using interpretive criteria delineated in the assay protocol.

*Influenza detection by RT-PCR*

Total nucleic acid extract from 100 microliters of upper respiratory specimen collected in viral transport medium was prepared using the MagNA Pure LC Total Nucleic Acid Isolation Kit and MagNA Pure 96 automated extraction platform or QIAamp 96 Virus QiaCube HT kit and QiaCube HT automated extraction system. Extracts (100 microliters eluate volume) were tested by RT-PCR using the QuantStudio 3, QuantStudio 5, or QuantStudio 6 Real-Time PCR System for influenza A and B using the *CDC Human Influenza Virus Real-Time RT-PCR Diagnostic Panel, Influenza A/B Typing Kit (VER 2)* with Superscript III Platinum One-Step Quantitative RT-PCR System containing ROX passive reference dye (Invitrogen, Waltham, MA). Subtyping of influenza A-positive specimens by RT-PCR was performed using the *CDC Human Influenza Virus Real-Time RT-PCR Diagnostic Panel, Influenza A Subtyping Kit (VER 3)* with Superscript III Platinum One-Step Quantitative RT-PCR System containing ROX passive reference dye. Each specimen also was tested for RNP using TaqPath 1-Step RT-qPCR Master Mix CG (<https://www.fda.gov/media/134922/download>). PCR reactions consisted of 45 amplification cycles, and Ct values of any magnitude were deemed positive when represented by a characteristic specific amplification curve. A valid influenza A subtype was contingent on co-detection of the universal influenza type A sequence target. Absence of influenza A and/or B detection in specimens registering RNP Ct values ≥38 was considered inconclusive for the undetected virus(es).

*RSV detection by RT-PCR*

Total nucleic acid extract from 100 microliters of upper respiratory specimen collected in viral transport medium was prepared using the MagNA Pure LC Total Nucleic Acid Isolation Kit and MagNA Pure 96 automated extraction platform or QIAamp 96 Virus QiaCube HT kit and QiaCube HT automated extraction system. Extracts (100 microliters eluate volume) were tested by RT-PCR using the QuantStudio 3, QuantStudio 5, or QuantStudio 6 Real-Time PCR System for RSV (universal), RSV-A, and RSV-B.

A conserved sequence target in the RSV matrix gene was amplified using methods adapted from published procedures ([https://doi.org/10.1016/j.jviromet.2019.113676](https://nam12.safelinks.protection.outlook.com/?url=https%3A%2F%2Fdoi.org%2F10.1016%2Fj.jviromet.2019.113676&data=05%7C02%7Cjim.chappell%40vumc.org%7C27add10cef0f423f4b9d08dd9a228543%7Cef57503014244ed8b83c12c533d879ab%7C0%7C0%7C638836196020181678%7CUnknown%7CTWFpbGZsb3d8eyJFbXB0eU1hcGkiOnRydWUsIlYiOiIwLjAuMDAwMCIsIlAiOiJXaW4zMiIsIkFOIjoiTWFpbCIsIldUIjoyfQ%3D%3D%7C0%7C%7C%7C&sdata=%2FsJuL3ii587eZDVfhJiUoefi6X6V45uazS%2BlXrPEYB4%3D&reserved=0)) and unpublished protocols developed at the CDC. Pan-RSV screening employed AgPath-ID One-Step RT-PCR Reagents (Applied Biosystems), forward primer GGCAAATATGGAAACATACGTGAA (ThermoFisher, Waltham, MA), unpublished CDC reverse primer modified from ([https://doi.org/10.1016/j.jviromet.2019.113676](https://nam12.safelinks.protection.outlook.com/?url=https%3A%2F%2Fdoi.org%2F10.1016%2Fj.jviromet.2019.113676&data=05%7C02%7Cjim.chappell%40vumc.org%7C27add10cef0f423f4b9d08dd9a228543%7Cef57503014244ed8b83c12c533d879ab%7C0%7C0%7C638836196020181678%7CUnknown%7CTWFpbGZsb3d8eyJFbXB0eU1hcGkiOnRydWUsIlYiOiIwLjAuMDAwMCIsIlAiOiJXaW4zMiIsIkFOIjoiTWFpbCIsIldUIjoyfQ%3D%3D%7C0%7C%7C%7C&sdata=%2FsJuL3ii587eZDVfhJiUoefi6X6V45uazS%2BlXrPEYB4%3D&reserved=0)) (ThermoFisher), and unpublished CDC fluorescent hydrolysis probe modified from ([https://doi.org/10.1016/j.jviromet.2019.113676](https://nam12.safelinks.protection.outlook.com/?url=https%3A%2F%2Fdoi.org%2F10.1016%2Fj.jviromet.2019.113676&data=05%7C02%7Cjim.chappell%40vumc.org%7C27add10cef0f423f4b9d08dd9a228543%7Cef57503014244ed8b83c12c533d879ab%7C0%7C0%7C638836196020181678%7CUnknown%7CTWFpbGZsb3d8eyJFbXB0eU1hcGkiOnRydWUsIlYiOiIwLjAuMDAwMCIsIlAiOiJXaW4zMiIsIkFOIjoiTWFpbCIsIldUIjoyfQ%3D%3D%7C0%7C%7C%7C&sdata=%2FsJuL3ii587eZDVfhJiUoefi6X6V45uazS%2BlXrPEYB4%3D&reserved=0)) (Integrated DNA Technologies, Coralville, IA). RT-PCR reactions contained 12.5 microliters 2x reaction mix, 1.0 micromolar each forward and reverse primer, 0.25 micromolar probe, 1.0 microliter enzyme, and 5 microliters nucleic acid extract in a total volume of 25 microliters nuclease-free H_2_O. Cycling conditions consisted of 45^o^C x 10 min, 95^o^C x 10 min, and 45 cycles of 95^o^C x 15 sec followed by 55^o^C x 60 sec. Ct values of any magnitude were deemed positive when represented by a characteristic specific amplification curve.

Subgroup differentiation of RSV screen-positive specimens employed a CDC method using AgPath-ID One-Step RT-PCR Reagents, unpublished CDC nucleocapsid (N) gene forward primer modified from ([https://doi.org/10.1016/j.jviromet.2019.113676](https://nam12.safelinks.protection.outlook.com/?url=https%3A%2F%2Fdoi.org%2F10.1016%2Fj.jviromet.2019.113676&data=05%7C02%7Cjim.chappell%40vumc.org%7C27add10cef0f423f4b9d08dd9a228543%7Cef57503014244ed8b83c12c533d879ab%7C0%7C0%7C638836196020181678%7CUnknown%7CTWFpbGZsb3d8eyJFbXB0eU1hcGkiOnRydWUsIlYiOiIwLjAuMDAwMCIsIlAiOiJXaW4zMiIsIkFOIjoiTWFpbCIsIldUIjoyfQ%3D%3D%7C0%7C%7C%7C&sdata=%2FsJuL3ii587eZDVfhJiUoefi6X6V45uazS%2BlXrPEYB4%3D&reserved=0)) (ThermoFisher), unpublished CDC N gene reverse primer modified from ([https://doi.org/10.1016/j.jviromet.2019.113676](https://nam12.safelinks.protection.outlook.com/?url=https%3A%2F%2Fdoi.org%2F10.1016%2Fj.jviromet.2019.113676&data=05%7C02%7Cjim.chappell%40vumc.org%7C27add10cef0f423f4b9d08dd9a228543%7Cef57503014244ed8b83c12c533d879ab%7C0%7C0%7C638836196020181678%7CUnknown%7CTWFpbGZsb3d8eyJFbXB0eU1hcGkiOnRydWUsIlYiOiIwLjAuMDAwMCIsIlAiOiJXaW4zMiIsIkFOIjoiTWFpbCIsIldUIjoyfQ%3D%3D%7C0%7C%7C%7C&sdata=%2FsJuL3ii587eZDVfhJiUoefi6X6V45uazS%2BlXrPEYB4%3D&reserved=0)) (ThermoFisher), and RSV-A N-gene subgroup-specific fluorescent hydrolysis probe <FAM>ACACTCAACAAAGA<BHQ1dT>CAACTTCTRTCATCCAGCA-phosphate ([https://doi.org/10.1016/j.jviromet.2019.113676](https://nam12.safelinks.protection.outlook.com/?url=https%3A%2F%2Fdoi.org%2F10.1016%2Fj.jviromet.2019.113676&data=05%7C02%7Cjim.chappell%40vumc.org%7C27add10cef0f423f4b9d08dd9a228543%7Cef57503014244ed8b83c12c533d879ab%7C0%7C0%7C638836196020181678%7CUnknown%7CTWFpbGZsb3d8eyJFbXB0eU1hcGkiOnRydWUsIlYiOiIwLjAuMDAwMCIsIlAiOiJXaW4zMiIsIkFOIjoiTWFpbCIsIldUIjoyfQ%3D%3D%7C0%7C%7C%7C&sdata=%2FsJuL3ii587eZDVfhJiUoefi6X6V45uazS%2BlXrPEYB4%3D&reserved=0)) (Biosearch Technologies, Novato, CA) or unpublished CDC RSV-B N-gene fluorescent hydrolysis probe modified from ([https://doi.org/10.1016/j.jviromet.2019.113676](https://nam12.safelinks.protection.outlook.com/?url=https%3A%2F%2Fdoi.org%2F10.1016%2Fj.jviromet.2019.113676&data=05%7C02%7Cjim.chappell%40vumc.org%7C27add10cef0f423f4b9d08dd9a228543%7Cef57503014244ed8b83c12c533d879ab%7C0%7C0%7C638836196020181678%7CUnknown%7CTWFpbGZsb3d8eyJFbXB0eU1hcGkiOnRydWUsIlYiOiIwLjAuMDAwMCIsIlAiOiJXaW4zMiIsIkFOIjoiTWFpbCIsIldUIjoyfQ%3D%3D%7C0%7C%7C%7C&sdata=%2FsJuL3ii587eZDVfhJiUoefi6X6V45uazS%2BlXrPEYB4%3D&reserved=0)) (Biosearch). RT-PCR reactions contained 12.5 microliters 2x reaction mix, 1.0 micromolar each forward and reverse primer, 0.25 micromolar probe, 1.0 microliter enzyme, and 5 microliters nucleic acid extract in a total volume of 25 microliters nuclease-free H_2_O. Cycling conditions consisted of 45^o^C x 10 min, 95^o^C x 10 min, and 45 cycles of 95^o^C x 15 sec followed by 55^o^C x 60 sec. Ct values of any magnitude were deemed positive when represented by a characteristic specific amplification curve.

Each specimen also was tested for RNP using TaqPath 1-Step RT-qPCR Master Mix CG (<https://www.fda.gov/media/134922/download>). A valid RSV subgroup identification was contingent on co-detection of the universal RSV target. Absence of RSV detection in specimens registering RNP Ct values ≥40 was considered inconclusive for RSV RNA. All RT-PCR reactions were performed in single-plex format.

*HMPV detection by RT-PCR*

Total nucleic acid extract from 100 microliters of upper respiratory specimen collected in viral transport medium was prepared using the MagNA Pure LC Total Nucleic Acid Isolation Kit and MagNA Pure 96 automated extraction platform or QIAamp 96 Virus QiaCube HT kit and QiaCube HT automated extraction system. Extracts (100 microliters eluate volume) were tested by RT-PCR using the QuantStudio 3, QuantStudio 5, or QuantStudio 6 Real-Time PCR System for a conserved sequence target in the hMPV fusion gene (<https://doi.org/10.1128/jcm.02270-10)>—forward primer CAAGTGTGACATTGCTGAYCTRAA (Biosearch), reverse primer ACTGCC GCACAACATTTAGRAA (Biosearch), and fluorescent hydrolysis probe <FAM>TGGCYGTYAGCTTCAGTCAATTCAACAGA<BHQ-1> (Biosearch)—with AgPath-ID One-Step RT-PCR Reagents. RT-PCR reactions contained 12.5 microliters 2x reaction mix, 1.0 micromolar each forward and reverse primer, 0.25 micromolar probe, 1.0 microliter enzyme, and 5 microliters nucleic acid extract in a total volume of 25 microliters nuclease-free H_2_O. Cycling conditions consisted of 45^o^C x 10 min, 95^o^C x 10 min, and 45 cycles of 95^o^C x 15 sec followed by 55^o^C x 60 sec. Ct values of any magnitude were deemed positive when represented by a characteristic specific amplification curve.

Each specimen also was tested for RNP using TaqPath 1-Step RT-qPCR Master Mix CG (<https://www.fda.gov/media/134922/download>). Absence of hMPV detection in specimens registering RNP Ct values ≥40 was considered inconclusive for hMPV RNA.

**3. Analytic methods**

*Defining immunocompromising conditions*

Immunocompromising conditions included active solid organ or hematologic cancer (defined as newly diagnosed cancer or cancer treatment within the past 6 months); solid organ transplant; bone marrow/stem cell transplant; HIV infection; congenital immunodeficiency syndrome; use of an immunosuppressive medication within the past 30 days; splenectomy; or another condition that causes moderate or severe immunosuppression.

*Bootstrap-based P-values and 95% CIs for differences in VE*

We used a non-parametric bootstrapping method to formally assess statistical evidence for differences in VE between in-hospital severe outcomes and hospitalization. Overlapping 95% CIs have been sometimes used as a heuristic to assess differences in VE, but are known to be conservative and do not account for correlation between effect estimates [1]. A bootstrap-based method should better account for the implicit correlation between effect estimates that may arise because in-hospital severe outcomes are nested within hospitalization overall.

Across 10,000 iterations separately for adults ≥18 and ≥65 years, we use the following approach:

1. Draw a bootstrapped dataset with size equal to the original dataset using case-resampling, i.e. sample rows of the original dataset with replacement.
2. Fit separate logistic regression models for the association of 2024–2025 COVID-19 vaccination with COVID-19–associated hospitalization and for each of the severe in-hospital outcomes. Letting $\beta_{1}$ represent the association of vaccination with case-control status on the logit scale and $\phi$ is a vector of coefficients corresponding to the confounders included in the model, including age, sex, race and ethnicity, geographic region (U.S. Department of Health and Human Services Region), calendar time (biweekly intervals), and Charlson comorbidity index:

$$logit\left( case status \right)= \beta_{0}+ \beta_{1}*vaccination+\phi*confounders$$

$VE_{1}$ representing VE against COVID-19–associated hospitalization is therefore given by $100 - 100*exp(\beta_{1})$. Similarly, the association of vaccination with case-control status restricting to COVID-19 cases with a given severe in-hospital outcome is given by $\beta_{2}$:

$$logit\left( case status \right)= \beta_{0}+ \beta_{2}*vaccination+\phi*confounders$$

And $VE_{2}$ representing VE against the COVID-19–associated severe in-hospital outcome is given by $100 - 100*exp(\beta_{2})$. Because the number of cases with 2024–2025 vaccination experiencing IMV receipt or death was low (n = 6), in some bootstrap samples, no vaccinated cases with this outcome were selected causing convergence problems for standard logistic regression. In such cases, we used Firth regression instead to estimate $\beta_{2}$.

3. Calculate the differences $\beta_{2}^{*}- \beta_{1}^{*}$ and $VE_{1}^{*} - VE_{2}^{*}$, where * indicates these quantities were derived from bootstrapping. Store the results and proceed to the next iteration.

Using the bootstrapped distribution of $\beta_{2}^{*}- \beta_{1}^{*}$, we calculated *P*-values by inverting confidence intervals [2]. Under this approach, a series of confidence intervals were calculated at the $1-\alpha$ level for a range of $\alpha$ values, with the *P*-value defined as the smallest $\alpha$ such that 0 is not contained within the CI for $\beta_{2}^{*}- \beta_{1}^{*}$. We used bias-corrected and accelerated bootstrap intervals to calculate CIs adjusting for bias and skewness in the bootstrap distribution [3].

In a sensitivity analysis, we compared results with a different approach used to calculate bootstrap-based *P-*values as described in Fox (2008) [4]. We used the bootstrapped distribution $\beta_{2}^{*}- \beta_{1}^{*}$ and shifted it by the observed difference in coefficients $\hat{\beta_{1}}-\hat{\beta_{2}}$ in the original dataset to approximate a null distribution around the null hypothesis of ${H_{0}:\beta}_{2}-\beta_{1} = 0$ (see also [5,6]). This method assumed translation invariance of these two distributions. We then calculated the proportion of shifted $\left| \left( \beta_{2}^{*}-\beta_{1}^{*} \right)-\left( \hat{\beta_{1}}-\hat{\beta_{2}} \right) \right|$ values that were greater in magnitude than $\left| \hat{\beta_{1}}-\hat{\beta_{2}} \right|$.

The two approaches relied on different assumptions but appeared to yield generally similar *P*-values (Supplementary Table 1). We also computed and presented 95% CIs for the difference in VE on the percentage scale to help interpret effect sizes.

**Supplementary Table 1. Bootstrap-based^a^ *P-*values and 95% confidence intervals for differences in the effectiveness of 2024–2025 COVID-19 vaccine against COVID-19–associated hospitalization versus in-hospital severe outcomes – IVY Network, 26 hospitals, September 1, 2024–April 30, 2025.**

| Age group and outcomes being compared | *P*-value obtained by inverting confidence intervals | *P*-value obtained by shifting bootstrap distribution to generate a null distribution | 95% CI for percentage difference in VE against severe outcome minus VE against hospitalization |
| --- | --- | --- | --- |
| ≥18 years |  |  |  |
| Supplemental oxygen therapy  versus hospitalization | 0.142 | 0.150 | (-2%, 15%) |
| Acute respiratory failure versus  hospitalization | 0.413 | 0.405 | (-14%, 28%) |
| ICU admission versus  hospitalization | 0.074 | 0.094 | (-2%, 38%) |
| IMV or death versus  hospitalization^b^ | 0.004 | 0.043 | (18%, 57%) |
| ≥65 years |  |  |  |
| Supplemental oxygen therapy  versus hospitalization | 0.458 | 0.465 | (-5%, 12%) |
| Acute respiratory failure versus  hospitalization | 0.679 | 0.670 | (-35%, 16%) |
| ICU admission versus  hospitalization | 0.420 | 0.391 | (-17%, 31%) |
| IMV or death versus  hospitalization | 0.093 | 0.116 | (-3%, 47%) |

^a^ To test the hypothesis that VE differed by outcome, *P-*values and 95% CIs for differences in VE against hospitalization versus VE against severe outcomes were calculated using bootstrapping with 10,000 replicates. In each iteration, the difference in regression coefficients between a severe outcome and hospitalization was computed. We used Firth regression in rare iterations when no vaccinated cases experiencing IMV receipt or death (n = 6) were sampled, which caused convergence problems for standard logistic regression. We calculated *P*-values using the bootstrap replicates by inverting confidence intervals [2] and by a method described by Fox for approximating the null distribution (2008) [4].

^b^ indicates *P*-value for difference in regression coefficients is < 0.05 using both methods.

**Supplementary Table 2. Counts of spike protein N-terminal domain substitutions or deletions by Nextstrain clade – IVY Network, 26 hospitals, September 1, 2024–April 30, 2025.**

| N-terminal domain substitution or deletion | Nextstrain clade |
| --- | --- |
| None (n = 62) | - 24C (KP.3) [n = 37] - 24B (JN.1.11.1) [n = 11] - 24A (JN.1) [n = 8] - 25C (XFG) [n = 3] - 24D (XDV.1) [n = 2] - 24E (KP.3.1.1) [n = 1] |
| T22N and F59S substitutions (n = 237) | - 24F (XEC) [n = 217] - recombinant [n = 17] - 24A (JN.1) [n = 1] - 24B (JN.1.11.1) [n = 1] - 24D (XDV.1) [n = 1] |
| T22N substitution^a^ (n = 290) | - 24F (XEC) [n = 217] - 24H (LF.7) [n = 28] - recombinant [n = 21] - 24A (JN.1) [n = 12] - 25A (LP.8.1) [n = 4] - 24C (KP.3) [n = 3] - 24B (JN.1.11.1) [n = 2] - 24D (XDV.1) [n = 1] - 24E (KP.3.1.1) [n = 1] - 24G (KP.2.3) [n = 1] |
| S31 deletion^a^ (n = 602) | - 24E (KP.3.1.1) [n = 347] - 25A (LP.8.1) [n = 134] - 24G (KP.2.3) [n = 43] - 24A (JN.1) [n = 41] - 24B (JN.1.11.1) [n = 21] - recombinant [n = 8] - 24C (KP.3) [n = 7] - 24F (XEC) [n = 1] |

^a^ includes n = 9 SARS-CoV-2 strains with both the S31 deletion and T22N substitution.

**Supplementary Figure 1. Participant flow diagram and analytic cohorts.**

**11,595** patients admitted during

September 1, 2024–April 30, 2025 with COVID-19–like illness

**3,102** Excluded^a^:

• **1,948** Controls received a positive or indeterminate influenza test result or influenza testing was not done

**• 521** Controls aged ≥60 years who received a positive

RSV test result

• **186** SARS-CoV-2 testing indeterminate or not done

• **147** Cases received a positive or indeterminate influenza
 test result

• **93** Did not meet eligibility criteria

• **80** Cases received a positive or indeterminate RSV test result

• **60** Received 2024–2025 COVID-19 vaccine dose <7 days before

illness onset

• **43** Withdrew

• **16** Received more than one 2024–2025 vaccine dose

• **11** Cases received a positive or indeterminate hMPV test result

• **6** Unknown vaccination status

**• 5** Unknown or missing covariates

**• 4** Unknown or missing immune status

**8,493** patients included in analysis

**6,605 Controls**

• **5,381** No 2024–2025 dose

• **1,224** Received 2024–2025 dose

**1,888 Cases**

• **1,672** No 2024–2025 dose

• **216** Received 2024–2025 dose

)

Vaccine effectiveness against hospitalization by time since dose and against severe in-hospital outcomes

Lineage- and mutation-specific vaccine effectiveness against hospitalization

**951 Sequenced cases**

• **846** No 2024–2025 dose

• **105** Received 2024–2025 dose

**^a^** Exclusions are not mutually exclusive.

**Supplementary Figure 2. Numbers of COVID-19 cases and test-negative controls by admission week – IVY Network, 26 hospitals, September 1, 2024–April 30, 2025**.

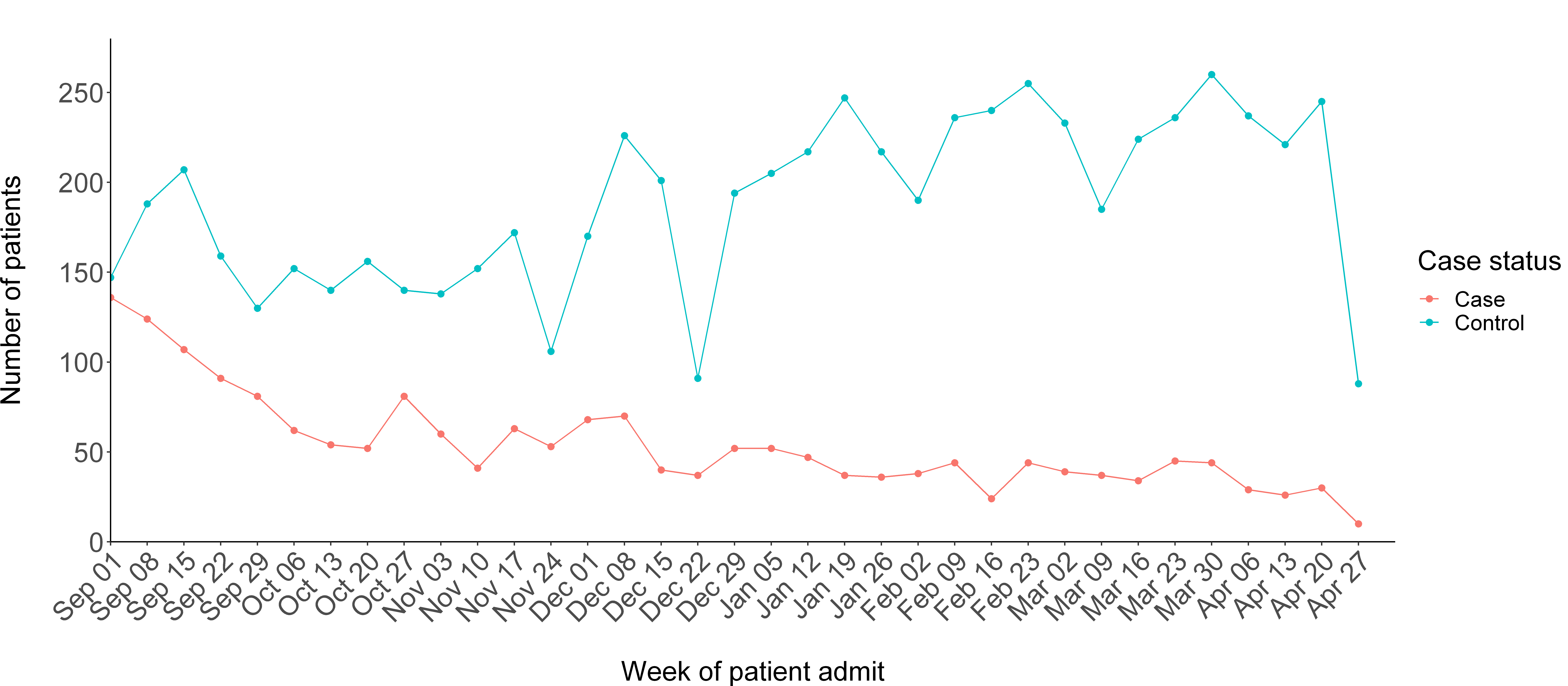

Dates are for the start of the admission week.

**Supplementary Figure 3. 2024–2025 COVID-19 vaccination coverage by admission week among test-negative controls – IVY Network, 26 hospitals, September 1, 2024–April 30, 2025**.^a^

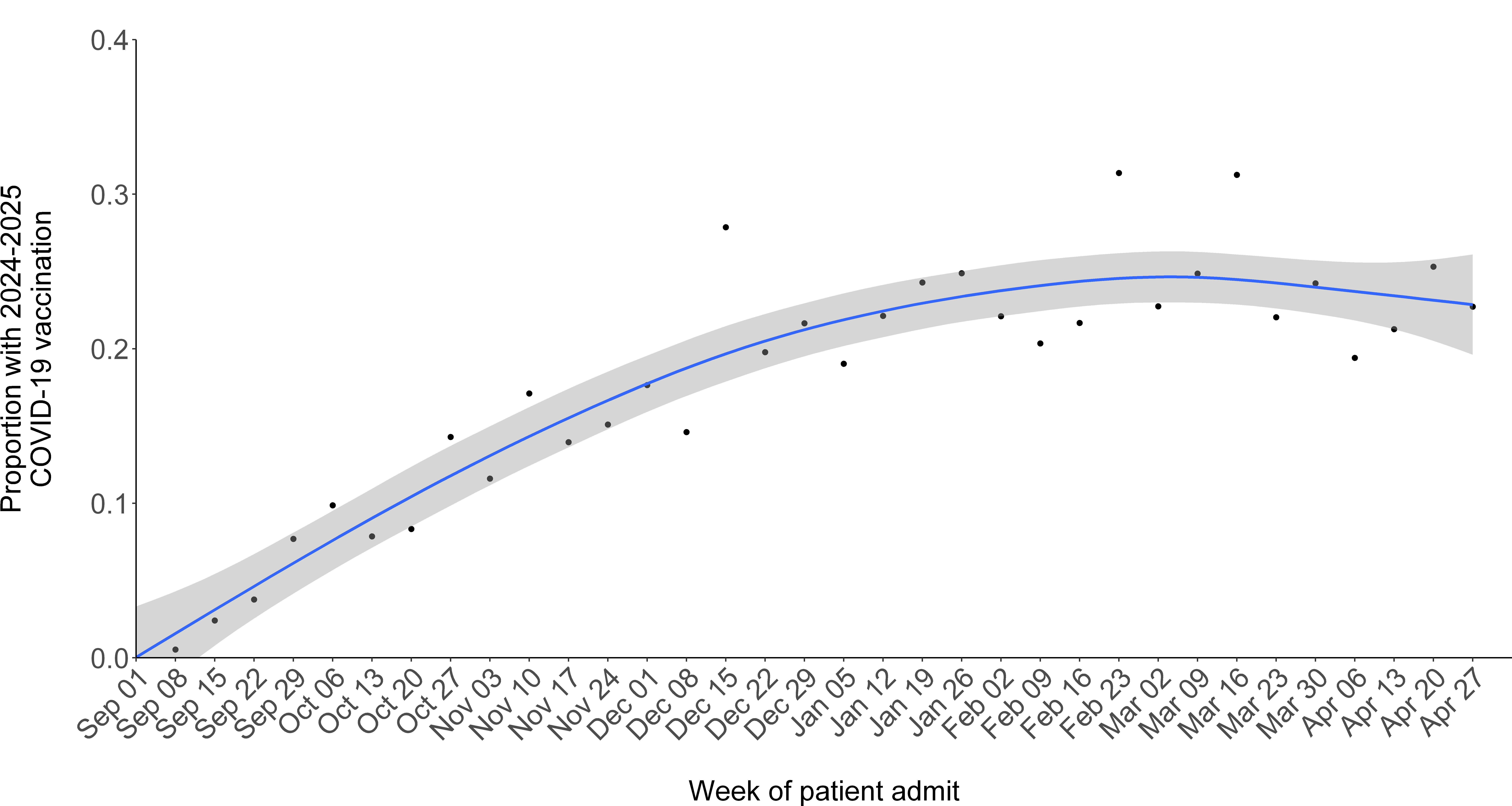

^a^ Locally estimated scatterplot smoothing (LOESS) was used to smooth the weekly data and generate 95% confidence intervals.

Dates are for the start of the admission week.

**Supplementary Figure 4.** **Effectiveness of 2024–2025 COVID-19 vaccine against COVID-19–associated hospitalization among immunocompetent adults by time since dose receipt (60-day windows) and age group – IVY Network, 26 hospitals, September 1, 2024–April 30, 2025.**

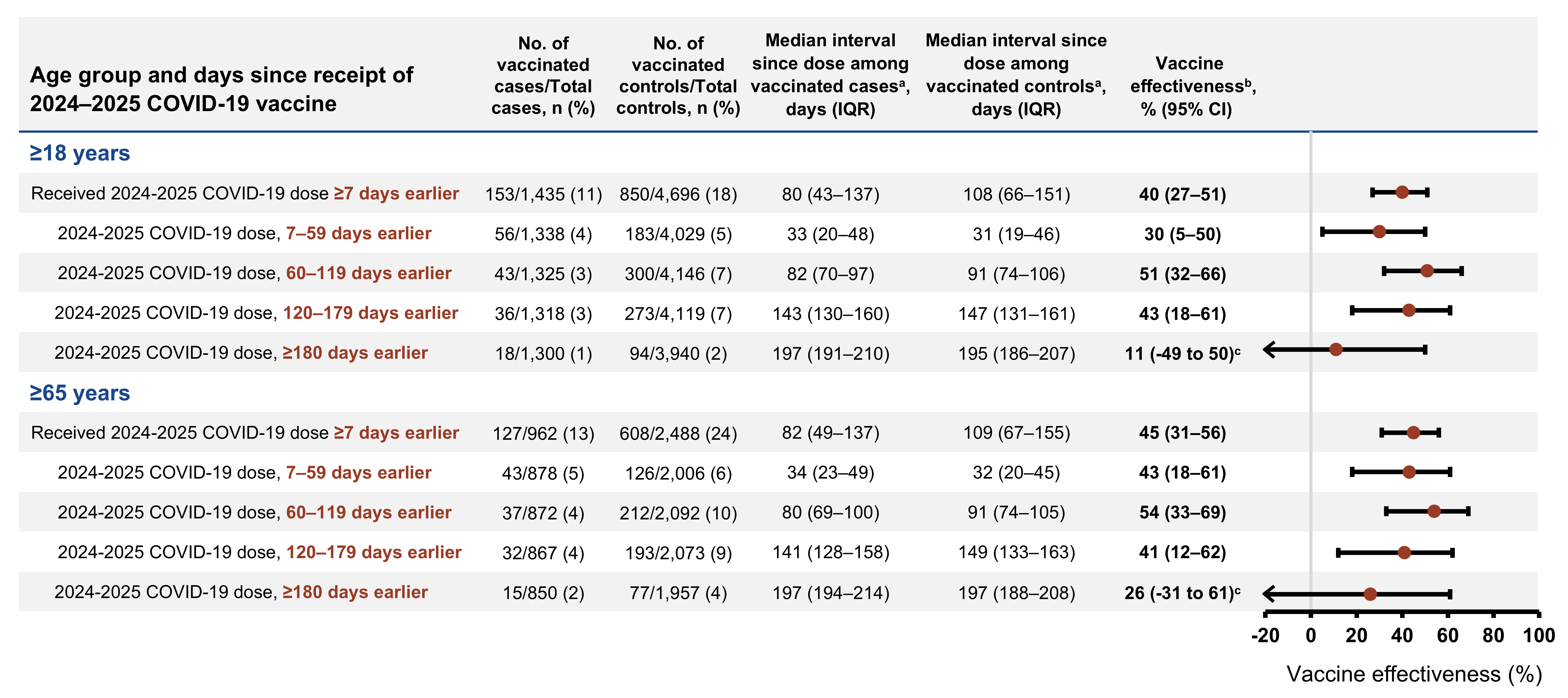

Abbreviations: CI, confidence interval; COVID-19, coronavirus disease 2019; IQR, interquartile range; IVY, Investigating Respiratory Viruses in the Acutely Ill; SARS-CoV-2, severe acute respiratory syndrome coronavirus 2.

^a^ Time since vaccination with a 2024–2025 COVID-19 vaccine.

^b^ Vaccine effectiveness was calculated by comparing the odds of 2024–2025 COVID-19 vaccination in cases and controls using the equation: (1 – adjusted odds ratio) x 100%. Odds ratios were estimated by multivariable logistic regression adjusted for age, sex, race and ethnicity, geographic region (U.S. Department of Health and Human Services Region), calendar time (biweekly intervals), and Charlson comorbidity index.

^c^ Estimates are imprecise due to limited numbers of enrolled patients with dose receipt ≥180 days earlier than hospitalization.

**Supplementary Figure 5.** **Effectiveness of 2024–2025 COVID-19 vaccine against COVID-19–associated hospitalization among immunocompromised^a^ adults by age group – IVY Network, 26 hospitals, September 1, 2024–April 30, 2025.**

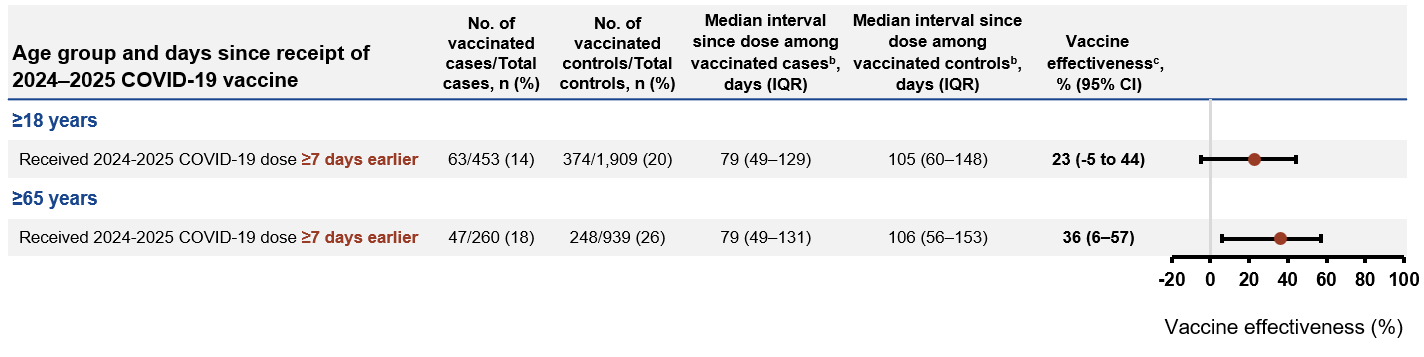

Abbreviations: CI, confidence interval; COVID-19, coronavirus disease 2019; IQR, interquartile range; IVY, Investigating Respiratory Viruses in the Acutely Ill; SARS-CoV-2, severe acute respiratory syndrome coronavirus 2.

^a^ Immunocompromising conditions included active solid tumor or hematologic malignancy (defined as newly diagnosed malignancy or treatment within the past 6 months); solid organ transplant; hematopoietic cell transplant; HIV infection; congenital immunodeficiency syndrome; use of an immunosuppressive medication within the past 30 days; splenectomy; or another condition that causes moderate or severe immunosuppression.

^b^ Time since vaccination with a 2024–2025 COVID-19 vaccine.

^c^ Vaccine effectiveness was calculated by comparing the odds of 2024–2025 COVID-19 vaccination in cases and controls using the equation: (1 – adjusted odds ratio) x 100%. Odds ratios were estimated by multivariable logistic regression adjusted for age, sex, race and ethnicity, geographic region (U.S. Department of Health and Human Services Region), calendar time (biweekly intervals), and Charlson comorbidity index.

**Supplementary Figure 6. Effectiveness of 2024–2025 COVID-19 vaccine against COVID-19–associated hospitalization among cases with SARS-CoV-2 lineage successfully identified through whole-genome sequencing and among all cases by time since dose receipt – IVY Network, 26 hospitals, September 1, 2024–April 30, 2025.**

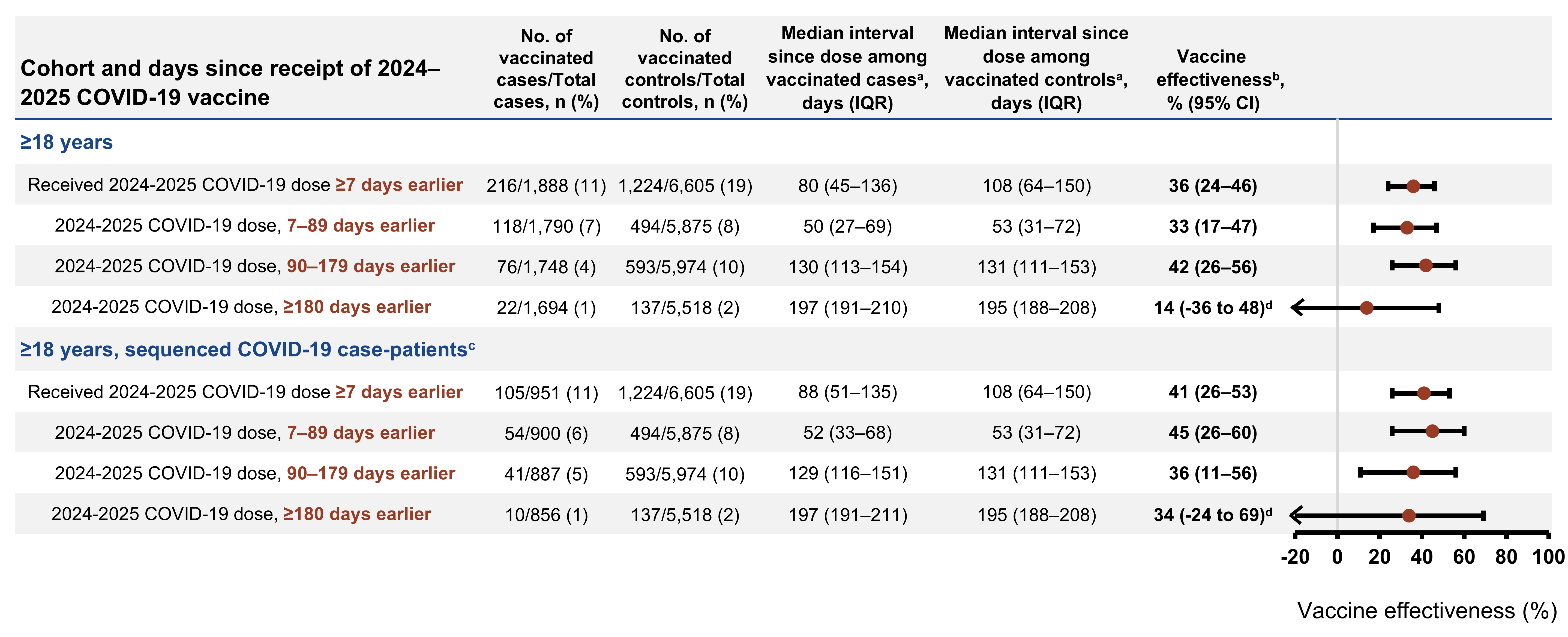

Abbreviations: CI, confidence interval; COVID-19, coronavirus disease 2019; IQR, interquartile range; IVY, Investigating Respiratory Viruses in the Acutely Ill; SARS-CoV-2, severe acute respiratory syndrome coronavirus 2.

^a^ Time since vaccination with a 2024–2025 COVID-19 vaccine.

^b^ Vaccine effectiveness was calculated by comparing the odds of 2024–2025 COVID-19 vaccination in cases and controls using the equation: (1 – adjusted odds ratio) x 100%. Odds ratios were estimated by multivariable logistic regression adjusted for age, sex, race and ethnicity, geographic region (U.S. Department of Health and Human Services Region), calendar time (biweekly intervals), and Charlson comorbidity index.

^c^ SARS-CoV-2 whole-genome sequences were considered adequate if they had a Nextclade completeness score greater than 80 and Nextclade quality control status of “good” or “mediocre.”

^d^ Estimates are imprecise due to limited numbers of enrolled patients with dose receipt ≥180 days earlier than hospitalization.

**Supplementary Figure 7. Number of COVID-19 cases by hospital admission week and SARS-CoV-2 spike protein N-terminal domain substitutions and deletions – IVY Network, 26 hospitals, September 1, 2024–April 30, 2025.^a^**

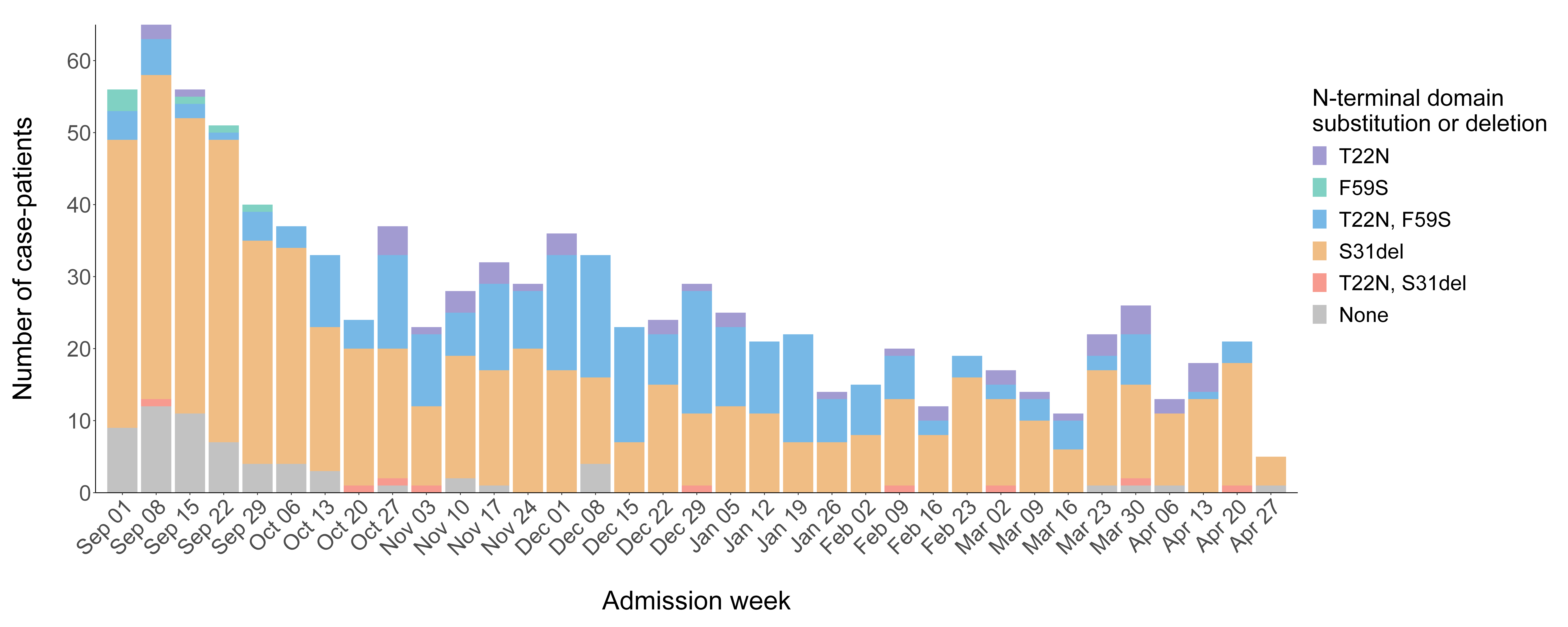

Abbreviations: COVID-19, coronavirus disease 2019; IVY, Investigating Respiratory Viruses in the Acutely Ill; SARS-CoV-2, severe acute respiratory syndrome coronavirus 2.

^a^ Dates are for the start of the admission week. SARS-CoV-2 spike protein substitutions and deletions were identified after conducting viral whole-genome sequencing.
